# Disparities in uptake of Shingrix^®^ vaccine in immunosuppressed individuals in England: a population-based cohort study

**DOI:** 10.1101/2025.08.27.25334559

**Authors:** Eleanor V.H. Barry, Anne M. Suffel, Jemma Walker, Nick Andrews, Colin Campbell, Rosalind Goudie, Simon de Lusignan, Meredith Leston, Sinéad M. Langan, Julia Stowe, Ian J. Douglas, Edward P.K. Parker, Kathryn E. Mansfield

**Affiliations:** Department for Infectious Disease Epidemiology and International Health, London School of Hygiene and Tropical Medicine; NIHR Health Protection Research Unit in Immunisation at the London School of Hygiene and Tropical Medicine; UK Health Security Agency; Nuffield Department of Primary Care Health Sciences, University of Oxford; Department of Non-communicable Disease Epidemiology, London School of Hygiene and Tropical Medicine; School of Health & Care Sciences, University of Lincoln

**Keywords:** Shingrix, immunosuppression, vaccine uptake, herpes zoster, shingles

## Abstract

**Background:** Herpes zoster (shingles) impacts health and quality of life, particularly in immunosuppressed individuals. The UK Shingrix^®^ zoster vaccination programme has been available for eligible immunosuppressed individuals since 2021. Understanding Shingrix^®^ uptake in immunosuppressed adults is critical for equitable vaccine delivery.

**Methods:** We conducted a population-based cohort study using primary care records from English practices contributing to Clinical Practice Research Datalink (CPRD) Aurum (1 September 2021-31 August 2023). We included immunosuppressed adults (70-79 years) with at least one year of prior registration. Our primary outcome was uptake of at least one dose of Shingrix^®^. Secondary outcomes were two-dose completion, inadvertent Zostavax^®^ receipt (live attenuated vaccine contraindicated in immunosuppression), and co-administration of Shingrix^®^ with seasonal influenza vaccine.

**Results:** We included 86,197 immunosuppressed adults, 17.3% received at least one Shingrix^®^ dose. Of those, 41.5% received two doses. Uptake was lower in older individuals, people from minority ethnic groups (e.g. Black vs. White OR 0.53, 95%CI 0.43-0.66), and the most vs. least deprived quintile (OR 0.61, 95%CI 0.56-0.67). While many factors showed strong relative differences, absolute differences were often modest. Absolute differences exceeded 5%, highlighting substantial disparity in vaccine coverage, for dementia (10.3% vs 17.5%), severe mental illness (10.8% vs 17.4%), Black ethnicity (10.0% vs 18.0%), care home residence (5.8% vs 17.5%), and deprivation (most 13.3% vs least 21.3%).

**Conclusions:** Our study highlights suboptimal Shingrix^®^ vaccine uptake during initial UK roll-out to immunosuppressed adults. Both relative and absolute differences reveal particularly low uptake among older people, those living with dementia or severe mental illness, and individuals from more deprived or minority ethnic backgrounds. Our results suggest a need for targeted strategies to improve access and reduce disparities in this vulnerable group.

**Highlights (to be submitted in separate file):**

– Shingrix^®^ uptake was 17.3% among eligible immunosuppressed adults in 2021-2023.
– Second-dose completion was at 41.5%.
– Uptake was lower in individuals who were older, more deprived, and of minority ethnicity.
– Uptake was lower in those with severe mental illness, dementia, and care home residence.

## Introduction

There are effective vaccines available to protect against herpes zoster (also known as shingles) and its complications.(1) Herpes zoster is a reactivation of the varicella-zoster virus (the virus causing chicken pox) that causes a painful rash.(2) Zoster impacts health and quality of life, and has a considerable economic burden.(3,4) The risk and severity of zoster is greater in those with immunosuppression.(5,6) Therefore, to help inform an effective zoster vaccination programme for eligible immunosuppressed individuals, it is vital that we understand the potential inequalities in access to zoster vaccination for individuals with immunosuppression (NB: we use the term immunosuppression throughout to refer to both immunocompromise [i.e., individuals with underlying health conditions causing immune system deficiency] and immunosuppression [i.e., individuals whose immune defences are weakened by medication or some other treatment]).

Since 2013, a live attenuated zoster vaccine (Zostavax^®^) has been available in the UK for individuals between 70 and 79 years of age.(7) The routine UK vaccination programme targets those aged 70 years on 1^st^ September of each year. Over the first nine years of the zoster vaccination programme, a phased catch-up programme ran to roll out vaccination to individuals aged 71-79 at the programme’s inception. Zostavax^®^, as a live vaccine, is contraindicated in those with immunosuppression/immunocompromise. Shingrix^®^ is a recombinant zoster vaccine that has been available in the UK since September 2021.(7) As Shingrix^®^ supply was initially limited, until September 2023, it was only available to individuals for whom the live Zostavax^®^ vaccine is contraindicated (i.e., those with severe immunosuppression).(7) From September 2023, Shingrix^®^ replaced Zostavax^®^ in the UK’s routine immunisation programme, and the programme expanded to all individuals aged 60-79, and immunosuppressed individuals aged 50 and over (with no upper age limit).(8)

We aimed to assess the performance of the current Shingrix^®^ zoster vaccination programme now that the Shingrix^®^ vaccination programme has been in place for eligible immunosuppressed individuals since 2021. We described Shingrix^®^ vaccine uptake in eligible immunosuppressed adults aged 70 to 79 following the introduction of the vaccine in September 2021. We also aimed to evaluate potential sociodemographic, clinical, and lifestyle-related disparities in Shingrix^®^ uptake in eligible immunosuppressed people.

## Methods

### Data sources

We used primary care records from the Clinical Practice Research Datalink (CPRD) Aurum database (1st September 2021 to 31st August 2023). CPRD Aurum includes de-identified data from participating general practices covering 24% of the UK population, and is broadly representative of the English population with respect to age, sex, ethnicity, and geographical region.(9)(10)

### Study population

We included all eligible immunosuppressed individuals aged 70 to 79 years between 2021 and the end of the study (i.e., born between 1942 and 1953), and with at least one year of prior registration with a CPRD Aurum practice during the study period (i.e., 1st September 2021 to 31st August 2023, when eligibility for the UK zoster vaccination programme changed).

To maintain anonymity, CPRD provides only year of birth for adults. We therefore assigned all participants a mid-year (1st July) date of birth. Individuals were eligible for inclusion from the latest of: study start (1^st^ September 2021), their 70th birthday, one year after their current practice registration (to ensure reliable capture of immunosuppression and baseline health status), or the date they met our immunosuppression definition. We excluded individuals who received Zostavax^®^ prior to cohort entry.

Individuals were eligible for follow up until the earliest of: end of study (i.e., 31^st^ August 2023), the middle of the year they turned 80, death, practice no longer contributing to CPRD, end of registration with practice, or based on predefined time periods (varying by reason for immunosuppression) after which individuals were no longer be considered immunosuppressed.

We defined immunosuppression using primary care morbidity coding and relevant prescriptions based on the definition of severe immunosuppression presented in the August 2021 version of the Shingles chapter of the Green Book (which provides UK guidelines on vaccines and vaccination procedures; **Supplementary Methods**).(7) We further categorised immunosuppression into higher and lower risk categories (and five constituent subcategories) based on recent evidence syntheses related to coronavirus disease 2019 (COVID-19) **(Table 1, Supplementary Table S1)**.(11,12)

**Table 1.**
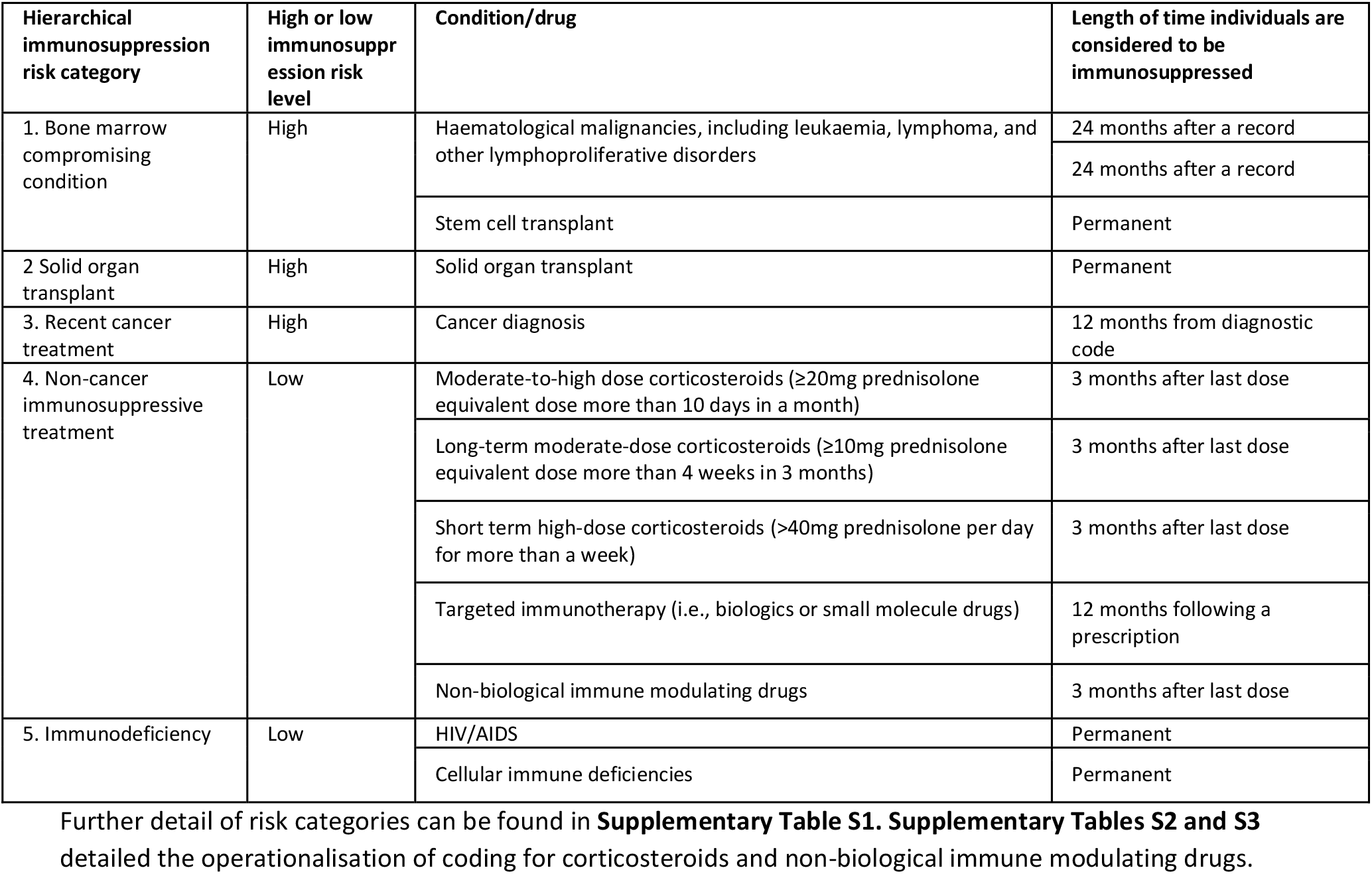
Summary of immunosuppression definition categories and risk levels.

| Hierarchical immunosuppression risk category | High or low immunosuppression risk level | Condition/drug | Length of time individuals are considered to be immunosuppressed |
| --- | --- | --- | --- |
| 1. Bone marrow compromising condition | High | Haematological malignancies, including leukaemia, lymphoma, and other lymphoproliferative disorders | 24 months after a record |
|  |  |  | 24 months after a record |
|  |  | Stem cell transplant | Permanent |
| 2 Solid organ transplant | High | Solid organ transplant | Permanent |
| 3. Recent cancer treatment | High | Cancer diagnosis | 12 months from diagnostic code |
| 4. Non-cancer immunosuppressive treatment | Low | Moderate-to-high dose corticosteroids (≥20mg prednisolone equivalent dose more than 10 days in a month) | 3 months after last dose |
|  |  | Long-term moderate-dose corticosteroids (≥10mg prednisolone equivalent dose more than 4 weeks in 3 months) | 3 months after last dose |
|  |  | Short term high-dose corticosteroids (>40mg prednisolone per day for more than a week) | 3 months after last dose |
|  |  | Targeted immunotherapy (i.e., biologics or small molecule drugs) | 12 months following a prescription |
|  |  | Non-biological immune modulating drugs | 3 months after last dose |
| 5. Immunodeficiency | Low | HIV/AIDS | Permanent |
|  |  | Cellular immune deficiencies | Permanent |

### Outcomes

We defined receipt of Shingrix^®^ vaccine using primary care morbidity coding and prescription records (as vaccination records may be recorded in either or both).(13,14) Our main outcome was receipt of at least one dose of Shingrix^®^ based on the first record of a relevant morbidity or prescription record for the vaccine.

We defined the first vaccine as the first recorded after entering the study (i.e., date when individuals became eligible). We categorised morbidity and prescription codes as: Shingrix^®^, Zostavax^®^, or ambiguous (i.e., records for zoster vaccination that could represent receipt of either Shingrix^®^ or Zostavax^®^). For each individual, we selected the first record indicating receipt of a zoster vaccine and any subsequent zoster vaccine records within the same week. Where there were multiple records we took the following approach to determine which vaccine product had been received: 1) Individuals with a record classified as Shingrix^®^, with or without any ambiguous records, were considered to have received Shingrix^®^; 2) individuals with a record for Zostavax^®^, with or without any ambiguous records, were considered to have received Zostavax^®^; 3) individuals with records for both Shingrix^®^ and Zostavax^®^, were classified as ambiguous; and 4) those with only ambiguous codes were classified as Shingrix^®^. We considered individuals recorded as receiving Shingrix^®^ or having an ambiguous record to have received Shingrix^®^ (on the basis that Shingrix^®^ is the recommended vaccine in this group).

We also considered second-dose completion as a secondary outcome. Guidelines recommend that the second Shingrix^®^ dose in immunosuppressed individuals should be received 8 weeks to 6 months after the first (8). Given that scheduling appointments to exactly follow these guidelines may not be possible, we allowed buffers on either side of the guideline recommended second dose timing. We therefore defined second-dose completion as any individual with a record indicating a second vaccination between 7 weeks and 13 months after their initial vaccination (13 months chosen as some individuals will be identified as not having a second dose at the next annual call-recall opportunity). We also reported the number of doses given outside this timeframe. We restricted the denominator population for second-dose completion to include individuals with eligible follow up for at least 13 months after first vaccination.

### Exposures and other variables

#### Potential uptake-related factors

We investigated potential sociodemographic, clinical, and lifestyle factors that may be associated with disparities in Shingrix^®^ vaccine uptake. All potential uptake-related factors were determined based on records before study entry. Descriptions of all variables and the morbidity code lists used to define them are provided in our online repository.(15)

Sociodemographic factors included age, sex, ethnicity, deprivation, geographic region, care home residence, and living alone. Age was recorded in one-year increments for individuals aged 70 to 79 years. Sex was classified as male or female. Ethnicity was based on morbidity coding in primary care records and categorised into five groups (White, South Asian, Black, and other or mixed, as used by the Office of National Statistics [ONS]).(16,17) Deprivation was categorised in quintiles of linked individual-level index of multiple deprivation (IMD).(9) We defined English geographic region using the ONS English regions based on the location of an individual’s primary care practice, categorised as: North East, North West, Yorkshire and the Humber, East Midlands, West Midlands, East of England, South West, South Central, London, South East coast. Living alone and care home residence were identified using primary care morbidity coding.(18,19)

We defined the underlying reason for immunosuppression using both primary care morbidity coding and prescribing records. Conditions were assigned to five immunosuppression-risk-level subcategories: 1) bone marrow compromising conditions; 2) solid organ transplant); 3) recent cancer treatment; 4) non-cancer immunosuppressive treatment; and 5) immunodeficiency. Subcategories 1-3 (bone marrow compromising, solid organ transplant, recent cancer treatment) were considered higher risk conditions. Subcategories 4 and 5 (non-cancer immunosuppressive treatment, and immunodeficiency) were considered lower-risk conditions (**Table 1, Supplementary Tables S2 and S3**). This binary classification was informed by research aiming to risk-stratify the heterogenous group of people with different forms of immunosuppression based on COVID-19-related health outcomes.(11,12) As individuals may have records in more than one immunosuppression-risk-level subcategory, we determined an individual’s immunosuppression level using hierarchical principles; if an individual had records, prior to start of eligibility for the vaccine, in more than one risk subcategory, they were classified at the highest of their risk levels. Similarly, if a code could potentially be assigned to more than one category, we classified it at the highest category (e.g., a solid organ transplant recipient on immunosuppressants was classified in the solid organ transplantation category rather than the non-cancer-related immunosuppressives category).

We defined the following clinical conditions based on a relevant morbidity code recorded in primary care records at any time before study entry: mental health conditions (common mental disorders [CMD] [i.e., depression, anxiety], severe mental illness [SMI] [schizophrenia, bipolar disorder, other psychoses]), dementia, autoimmune conditions (e.g., rheumatoid arthritis, inflammatory bowel disease), chronic obstructive pulmonary disease (COPD), and diabetes mellitus (DM). We defined chronic kidney disease (CKD) status based on estimated glomerular filtration rate calculated from serum creatinine test results recorded in primary care before study entry (**Supplementary Methods**). We included autoimmune conditions, COPD, CKD, and DM given their association with postherpetic neuralgia.(20)

We considered smoking status, being overweight/obese, and harmful alcohol use as potential uptake-related lifestyle factors. We pragmatically defined smoking status and being overweight/obese (i.e., body mass index [BMI] suggesting individuals were overweight or obese) based on primary care records for these measures, using the status recorded closest to study entry (**Supplementary Methods**).

#### Other variables

We defined recent shingles as any record indicating shingles in the year before individuals became eligible for Shingrix^®^ vaccination.

We defined co-administration of seasonal influenza vaccine with Shingrix^®^ as any record indicating receipt of influenza vaccine (recorded in primary care using morbidity codes or prescription records): 1) on the same date as that of the first recorded Shingrix^®^ vaccination; 2) within the 1-6 days either side; and 3) within the 7-14 days either side of Shingrix^®^ vaccination (to allow for the guideline-recommended 7-day interval between the two vaccinations).

### Statistical analyses

We initially described the characteristics of the study population. We then described the number and proportion of immunosuppressed individuals receiving: 1) at least one Shingrix^®^ vaccination (defined based on any record indicating vaccine receipt); and 2) two doses of Shingrix^®^ received 7 weeks to 13 months after the first (i.e., complete course).

We also described: 1) the number and proportion of eligible immunosuppressed individuals with a record indicating inadvertent Zostavax^®^; and 2) co-administration of influenza vaccination among Shingrix^®^ recipients (to highlight whether opportunistic co-administration of vaccines could offer a route to increased vaccine coverage).

We used multivariable logistic regression to estimate odds ratios (ORs) and 95% confidence intervals (CIs) for the association between each potential uptake-related factor and receipt of a first Shingrix^®^ vaccination. We used logistic regression given the potential misclassification of age-based vaccine eligibility due to only having year of birth and not precise date of birth (i.e., insufficient information to robustly define person-time eligible for vaccination).

Guidelines recommend that individuals with shingles should wait until their symptoms have resolved before receiving shingles immunisation.(7) However, since shingles will likely offer a natural boosting effect, the benefit of offering a zoster vaccine immediately following recovery is unclear. We therefore adjusted for recent shingles in analyses of potential uptake-related factors to account for recent shingles affecting the choice to vaccinate or not (e.g., because a clinician does not think it is worthwhile due to the natural boosting effect of shingles, or because an individual was offered vaccination while still experiencing the rash of shingles) and the potential for previous shingles to be related to the various potential factors associated with vaccine uptake.

We adjusted sequentially based on a hierarchical conceptual framework developed for each specific uptake-related factor of interest (13), adapting the modelling approach used in prior studies (justification of sequential modelling approach: **Supplementary Table S4**).(12,17) Model 1 adjusted for age, sex, and separately included each uptake-related factor (depending on the potential uptake-related factor under investigation). Model 2 additionally adjusted for recent shingles in the last year. Model 3 additionally adjusted for key sociodemographic factors including region, deprivation, and ethnicity. Model 4 additionally adjusted for immunosuppression risk level (high/low). Model 5 additionally adjusted for clinical factors (mental health conditions, dementia, DM, COPD, CKD). Model 6 (main analysis model) additionally adjusted for smoking, harmful alcohol use, and overweight/obese. Lastly, in a sensitivity analysis, Model 7 additionally adjusted for living alone and care home residence (we did not adjust these in our main analysis model as we felt they were unlikely to be captured reliably). We used a complete-case approach, excluding individuals with missing data on the variables included in a given model (missing data for: deprivation, ethnicity, overweight/obese, smoking).

We repeated our main analysis in a series of sensitivity analyses to assess the robustness of our findings (**Supplementary Table S5**).

In secondary analyses, we described uptake in each ethnic group within strata of IMD quintiles, and used likelihood ratio tests to investigate whether there was statistical evidence that deprivation modified the effect of ethnicity on Shingrix^®^ uptake.

### Software and reproducibility

We used R Version 4.3.2 for data management and analysis. Data management and statistical analysis code, and morbidity code lists used to define study variables are provided online.(15)

### Patient and public involvement

Our study design was supported by an online public involvement workshop including participants with immunosuppression in October 2023.

## Results

We included 86,197 adults aged 70-79 years with immunosuppression and active in CPRD between 1^st^ September 2021 and 31^st^ August 2023 (**Figure 1**). Median follow-up was 348 days (interquartile range [IQR] 104-729). Of the original 86,197, entry was highest on 1^st^ September 2021, when 50.4% (n=43,443) entered the study, with further peaks on 1^st^ July 2022 and 1^st^ July 2023 (**Supplementary Figure S1**). The cohort was skewed towards younger ages (e.g., 32.1% aged 70 years, 6.6% aged 79 years) and individuals were more likely to be women (56.9%) (**Table 2**). Ethnicity data were available for 95.7% (n=82,496) of the cohort, 88.2% (n=76,007/86,197) of the population were White. Forty-three percent (n=37,145) had higher-risk immunosuppressive conditions.

**Table 2.**
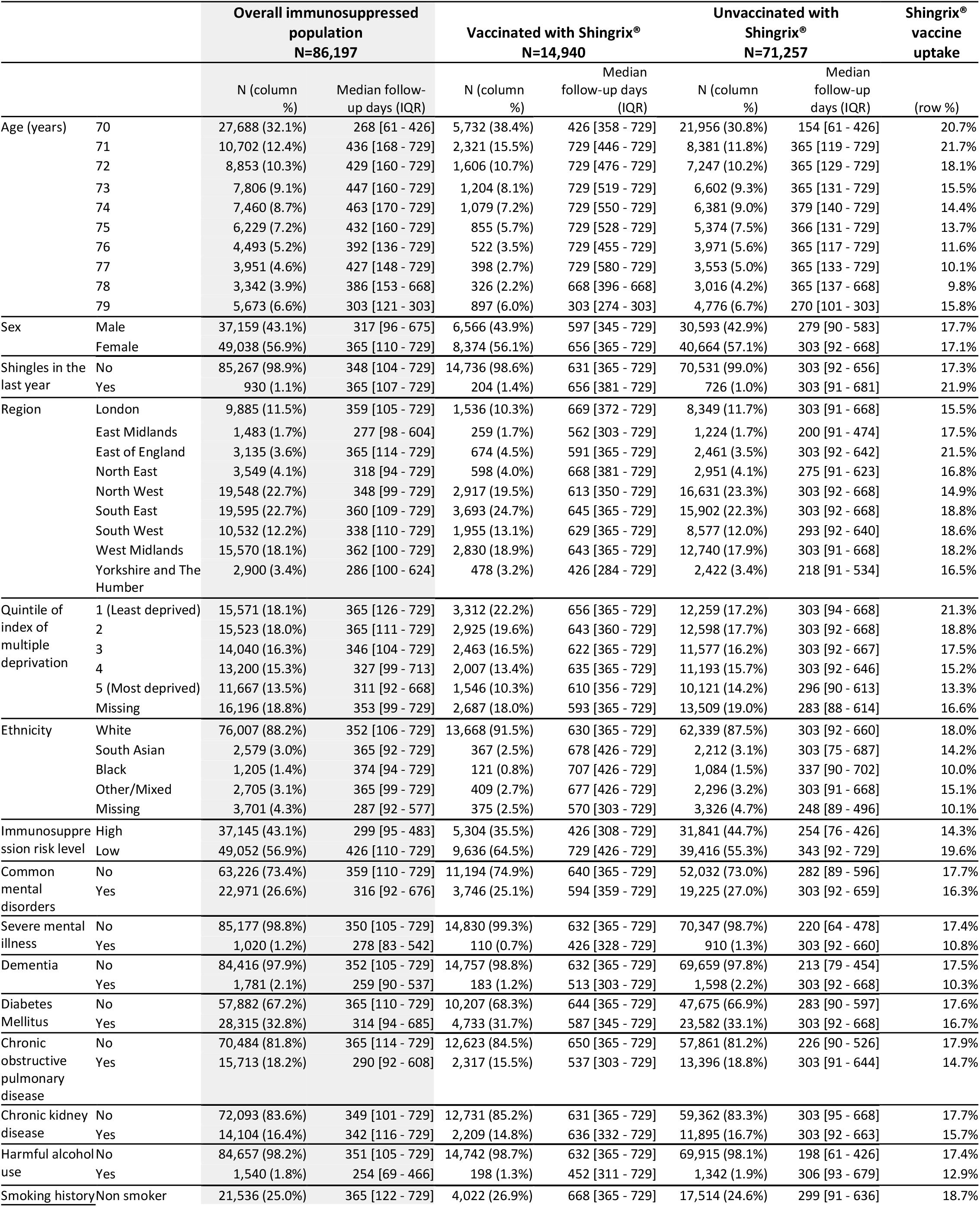
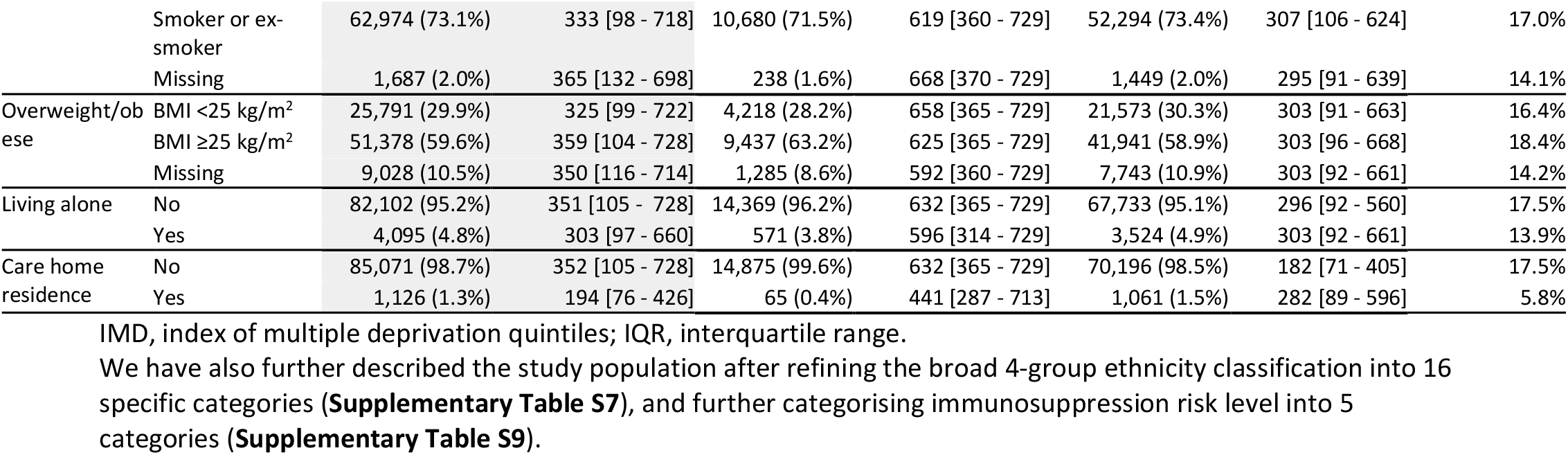
Baseline characteristics of the study cohort, and Shingrix(R) vaccine uptake (%).

|  |  | Overall immunosuppressed population<br>N=86,197 |  | Vaccinated with Shingrix®<br>N=14,940 |  | Unvaccinated with Shingrix®<br>N=71,257 |  | Shingrix® vaccine uptake |
| --- | --- | --- | --- | --- | --- | --- | --- | --- |
|  |  | N (column %) | Median follow-up days (IQR) | N (column %) | Median follow-up days (IQR) | N (column %) | Median follow-up days (IQR) | (row %) |
| Age (years) | 70 | 27,688 (32.1%) | 268 [61 - 426] | 5,732 (38.4%) | 426 [358 - 729] | 21,956 (30.8%) | 154 [61 - 426] | 20.7% |
|  | 71 | 10,702 (12.4%) | 436 [168 - 729] | 2,321 (15.5%) | 729 [446 - 729] | 8,381 (11.8%) | 365 [119 - 729] | 21.7% |
|  | 72 | 8,853 (10.3%) | 429 [160 - 729] | 1,606 (10.7%) | 729 [476 - 729] | 7,247 (10.2%) | 365 [129 - 729] | 18.1% |
|  | 73 | 7,806 (9.1%) | 447 [160 - 729] | 1,204 (8.1%) | 729 [519 - 729] | 6,602 (9.3%) | 365 [131 - 729] | 15.5% |
|  | 74 | 7,460 (8.7%) | 463 [170 - 729] | 1,079 (7.2%) | 729 [550 - 729] | 6,381 (9.0%) | 379 [140 - 729] | 14.4% |
|  | 75 | 6,229 (7.2%) | 432 [160 - 729] | 855 (5.7%) | 729 [528 - 729] | 5,374 (7.5%) | 366 [131 - 729] | 13.7% |
|  | 76 | 4,493 (5.2%) | 392 [136 - 729] | 522 (3.5%) | 729 [455 - 729] | 3,971 (5.6%) | 365 [117 - 729] | 11.6% |
|  | 77 | 3,951 (4.6%) | 427 [148 - 729] | 398 (2.7%) | 729 [580 - 729] | 3,553 (5.0%) | 365 [133 - 729] | 10.1% |
|  | 78 | 3,342 (3.9%) | 386 [153 - 668] | 326 (2.2%) | 668 [396 - 668] | 3,016 (4.2%) | 365 [137 - 668] | 9.8% |
|  | 79 | 5,673 (6.6%) | 303 [121 - 303] | 897 (6.0%) | 303 [274 - 303] | 4,776 (6.7%) | 270 [101 - 303] | 15.8% |
| Sex | Male | 37,159 (43.1%) | 317 [96 - 675] | 6,566 (43.9%) | 597 [345 - 729] | 30,593 (42.9%) | 279 [90 - 583] | 17.7% |
|  | Female | 49,038 (56.9%) | 365 [110 - 729] | 8,374 (56.1%) | 656 [365 - 729] | 40,664 (57.1%) | 303 [92 - 668] | 17.1% |
| Shingles in the last year | No | 85,267 (98.9%) | 348 [104 - 729] | 14,736 (98.6%) | 631 [365 - 729] | 70,531 (99.0%) | 303 [92 - 656] | 17.3% |
|  | Yes | 930 (1.1%) | 365 [107 - 729] | 204 (1.4%) | 656 [381 - 729] | 726 (1.0%) | 303 [91 - 681] | 21.9% |
| Region | London | 9,885 (11.5%) | 359 [105 - 729] | 1,536 (10.3%) | 669 [372 - 729] | 8,349 (11.7%) | 303 [91 - 668] | 15.5% |
|  | East Midlands | 1,483 (1.7%) | 277 [98 - 604] | 259 (1.7%) | 562 [303 - 729] | 1,224 (1.7%) | 200 [91 - 474] | 17.5% |
|  | East of England | 3,135 (3.6%) | 365 [114 - 729] | 674 (4.5%) | 591 [365 - 729] | 2,461 (3.5%) | 303 [92 - 642] | 21.5% |
|  | North East | 3,549 (4.1%) | 318 [94 - 729] | 598 (4.0%) | 668 [381 - 729] | 2,951 (4.1%) | 275 [91 - 623] | 16.8% |
|  | North West | 19,548 (22.7%) | 348 [99 - 729] | 2,917 (19.5%) | 613 [350 - 729] | 16,631 (23.3%) | 303 [92 - 668] | 14.9% |
|  | South East | 19,595 (22.7%) | 360 [109 - 729] | 3,693 (24.7%) | 645 [365 - 729] | 15,902 (22.3%) | 303 [92 - 668] | 18.8% |
|  | South West | 10,532 (12.2%) | 338 [110 - 729] | 1,955 (13.1%) | 629 [365 - 729] | 8,577 (12.0%) | 293 [92 - 640] | 18.6% |
|  | West Midlands | 15,570 (18.1%) | 362 [100 - 729] | 2,830 (18.9%) | 643 [365 - 729] | 12,740 (17.9%) | 303 [91 - 668] | 18.2% |
|  | Yorkshire and The Humber | 2,900 (3.4%) | 286 [100 - 624] | 478 (3.2%) | 426 [284 - 729] | 2,422 (3.4%) | 218 [91 - 534] | 16.5% |
| Quintile of index of multiple deprivation | 1 (Least deprived) | 15,571 (18.1%) | 365 [126 - 729] | 3,312 (22.2%) | 656 [365 - 729] | 12,259 (17.2%) | 303 [94 - 668] | 21.3% |
|  | 2 | 15,523 (18.0%) | 365 [111 - 729] | 2,925 (19.6%) | 643 [360 - 729] | 12,598 (17.7%) | 303 [92 - 668] | 18.8% |
|  | 3 | 14,040 (16.3%) | 346 [104 - 729] | 2,463 (16.5%) | 622 [365 - 729] | 11,577 (16.2%) | 303 [92 - 667] | 17.5% |
|  | 4 | 13,200 (15.3%) | 327 [99 - 713] | 2,007 (13.4%) | 635 [365 - 729] | 11,193 (15.7%) | 303 [92 - 646] | 15.2% |
|  | 5 (Most deprived) | 11,667 (13.5%) | 311 [92 - 668] | 1,546 (10.3%) | 610 [356 - 729] | 10,121 (14.2%) | 296 [90 - 613] | 13.3% |
|  | Missing | 16,196 (18.8%) | 353 [99 - 729] | 2,687 (18.0%) | 593 [365 - 729] | 13,509 (19.0%) | 283 [88 - 614] | 16.6% |
| Ethnicity | White | 76,007 (88.2%) | 352 [106 - 729] | 13,668 (91.5%) | 630 [365 - 729] | 62,339 (87.5%) | 303 [92 - 660] | 18.0% |
|  | South Asian | 2,579 (3.0%) | 365 [92 - 729] | 367 (2.5%) | 678 [426 - 729] | 2,212 (3.1%) | 303 [75 - 687] | 14.2% |
|  | Black | 1,205 (1.4%) | 374 [94 - 729] | 121 (0.8%) | 707 [426 - 729] | 1,084 (1.5%) | 337 [90 - 702] | 10.0% |
|  | Other/Mixed | 2,705 (3.1%) | 365 [99 - 729] | 409 (2.7%) | 677 [426 - 729] | 2,296 (3.2%) | 303 [91 - 668] | 15.1% |
|  | Missing | 3,701 (4.3%) | 287 [92 - 577] | 375 (2.5%) | 570 [303 - 729] | 3,326 (4.7%) | 248 [89 - 496] | 10.1% |
| Immunosuppression risk level | High | 37,145 (43.1%) | 299 [95 - 483] | 5,304 (35.5%) | 426 [308 - 729] | 31,841 (44.7%) | 254 [76 - 426] | 14.3% |
|  | Low | 49,052 (56.9%) | 426 [110 - 729] | 9,636 (64.5%) | 729 [426 - 729] | 39,416 (55.3%) | 343 [92 - 729] | 19.6% |
| Common mental disorders | No | 63,226 (73.4%) | 359 [110 - 729] | 11,194 (74.9%) | 640 [365 - 729] | 52,032 (73.0%) | 282 [89 - 596] | 17.7% |
|  | Yes | 22,971 (26.6%) | 316 [92 - 676] | 3,746 (25.1%) | 594 [359 - 729] | 19,225 (27.0%) | 303 [92 - 659] | 16.3% |
| Severe mental illness | No | 85,177 (98.8%) | 350 [105 - 729] | 14,830 (99.3%) | 632 [365 - 729] | 70,347 (98.7%) | 220 [64 - 478] | 17.4% |
|  | Yes | 1,020 (1.2%) | 278 [83 - 542] | 110 (0.7%) | 426 [328 - 729] | 910 (1.3%) | 303 [92 - 660] | 10.8% |
| Dementia | No | 84,416 (97.9%) | 352 [105 - 729] | 14,757 (98.8%) | 632 [365 - 729] | 69,659 (97.8%) | 213 [79 - 454] | 17.5% |
|  | Yes | 1,781 (2.1%) | 259 [90 - 537] | 183 (1.2%) | 513 [303 - 729] | 1,598 (2.2%) | 303 [92 - 668] | 10.3% |
| Diabetes Mellitus | No | 57,882 (67.2%) | 365 [110 - 729] | 10,207 (68.3%) | 644 [365 - 729] | 47,675 (66.9%) | 283 [90 - 597] | 17.6% |
|  | Yes | 28,315 (32.8%) | 314 [94 - 685] | 4,733 (31.7%) | 587 [345 - 729] | 23,582 (33.1%) | 303 [92 - 668] | 16.7% |
| Chronic obstructive pulmonary disease | No | 70,484 (81.8%) | 365 [114 - 729] | 12,623 (84.5%) | 650 [365 - 729] | 57,861 (81.2%) | 226 [90 - 526] | 17.9% |
|  | Yes | 15,713 (18.2%) | 290 [92 - 608] | 2,317 (15.5%) | 537 [303 - 729] | 13,396 (18.8%) | 303 [91 - 644] | 14.7% |
| Chronic kidney disease | No | 72,093 (83.6%) | 349 [101 - 729] | 12,731 (85.2%) | 631 [365 - 729] | 59,362 (83.3%) | 303 [95 - 668] | 17.7% |
|  | Yes | 14,104 (16.4%) | 342 [116 - 729] | 2,209 (14.8%) | 636 [332 - 729] | 11,895 (16.7%) | 303 [92 - 663] | 15.7% |
| Harmful alcohol use | No | 84,657 (98.2%) | 351 [105 - 729] | 14,742 (98.7%) | 632 [365 - 729] | 69,915 (98.1%) | 198 [61 - 426] | 17.4% |
|  | Yes | 1,540 (1.8%) | 254 [69 - 466] | 198 (1.3%) | 452 [311 - 729] | 1,342 (1.9%) | 306 [93 - 679] | 12.9% |
| Smoking history | Non smoker | 21,536 (25.0%) | 365 [122 - 729] | 4,022 (26.9%) | 668 [365 - 729] | 17,514 (24.6%) | 299 [91 - 636] | 18.7% |
|  | Smoker or ex-smoker | 62,974 (73.1%) | 333 [98 - 718] | 10,680 (71.5%) | 619 [360 - 729] | 52,294 (73.4%) | 307 [106 - 624] | 17.0% |
|  | Missing | 1,687 (2.0%) | 365 [132 - 698] | 238 (1.6%) | 668 [370 - 729] | 1,449 (2.0%) | 295 [91 - 639] | 14.1% |
| Overweight/obese | BMI <25 kg/m<sup>2</sup> | 25,791 (29.9%) | 325 [99 - 722] | 4,218 (28.2%) | 658 [365 - 729] | 21,573 (30.3%) | 303 [91 - 663] | 16.4% |
|  | BMI ≥25 kg/m<sup>2</sup> | 51,378 (59.6%) | 359 [104 - 728] | 9,437 (63.2%) | 625 [365 - 729] | 41,941 (58.9%) | 303 [96 - 668] | 18.4% |
|  | Missing | 9,028 (10.5%) | 350 [116 - 714] | 1,285 (8.6%) | 592 [360 - 729] | 7,743 (10.9%) | 303 [92 - 661] | 14.2% |
| Living alone | No | 82,102 (95.2%) | 351 [105 - 728] | 14,369 (96.2%) | 632 [365 - 729] | 67,733 (95.1%) | 296 [92 - 560] | 17.5% |
|  | Yes | 4,095 (4.8%) | 303 [97 - 660] | 571 (3.8%) | 596 [314 - 729] | 3,524 (4.9%) | 303 [92 - 661] | 13.9% |
| Care home residence | No | 85,071 (98.7%) | 352 [105 - 728] | 14,875 (99.6%) | 632 [365 - 729] | 70,196 (98.5%) | 182 [71 - 405] | 17.5% |
|  | Yes | 1,126 (1.3%) | 194 [76 - 426] | 65 (0.4%) | 441 [287 - 713] | 1,061 (1.5%) | 282 [89 - 596] | 5.8% |

IMD, index of multiple deprivation quintiles; IQR, interquartile range.

**Figure 1.**
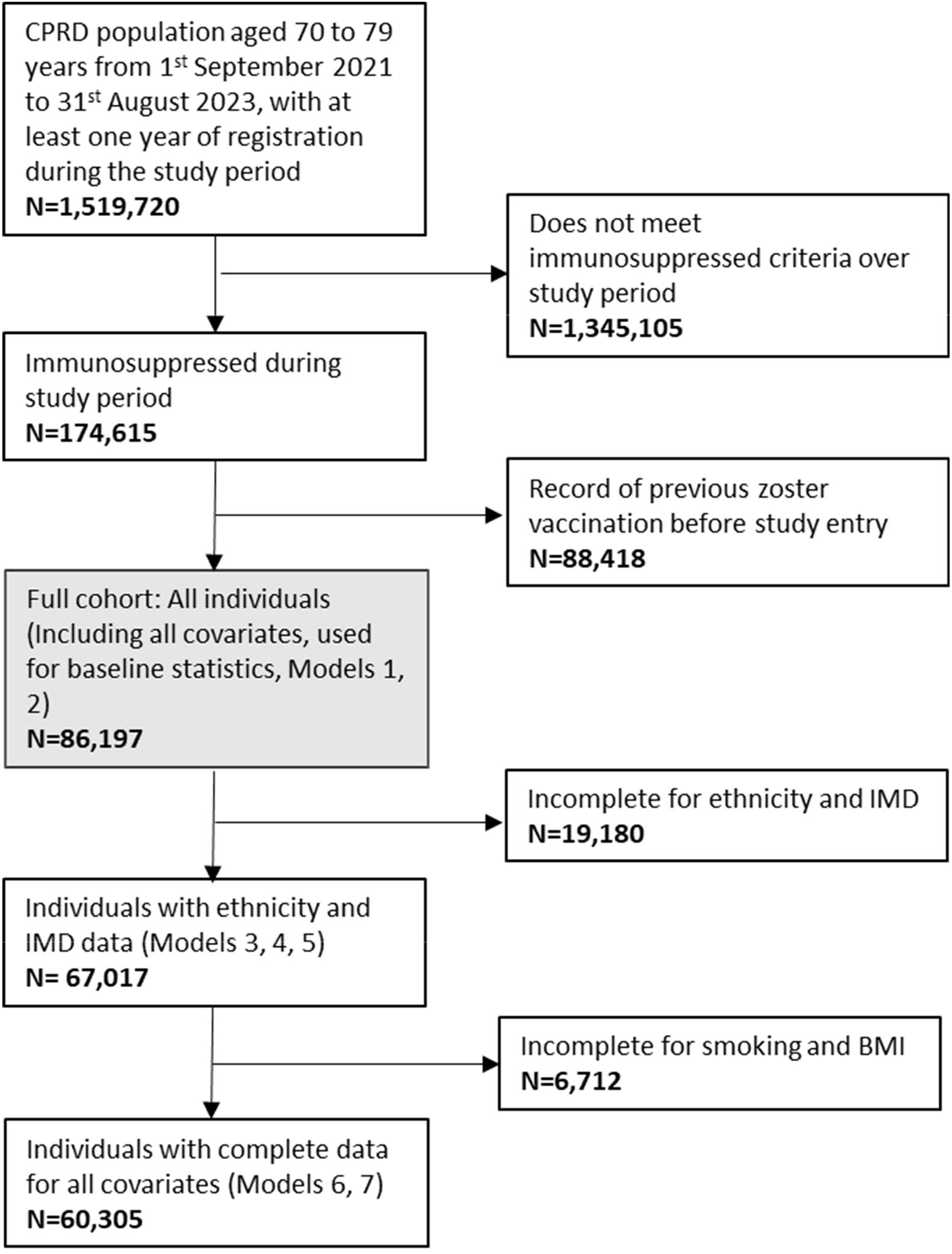
Study population flow chart. Where x represents each uptake-related factor: Model 1: age + sex (+ x) Model 2: Model 1 + prior shingles infection (+ x) Model 3: Model 2 + region + IMD + ethnicity (+x) Model 4: Model 3 + risk level (+ x), Model 5: Model 4 + CMD + SMI + dementia + DM + COPD + CKD (+ x) Model 6: Model 5 + harmful alcohol use + smoking + overweight/obese (+ x) Model 7: Model 6 + living alone + care home residence. See **Supplementary Table S4** for details. Abbreviations: BMI, body mass index; CPRD, Clinical Practice Research Datalink; IMD, index of multiple deprivation; CMD, Common mental disorder; SMI, Severe mental illness; SMI, severe mental illness; DM, diabetes mellitus; COPD, chronic obstructive pulmonary disease; CKD, chronic kidney disease.

### One-dose uptake

Uptake of at least one dose of Shingrix^®^ was 17.3% (14,940/86,197) in the overall population and 18.1% (10,924/60,305) in those with complete data (on deprivation, ethnicity, overweight/obese, smoking) and therefore included in the fully adjusted model. Of the 14,940 first doses recorded, only 5.8% (n=865) were Shingrix^®^ specific while 94.2% (n=14,075) were classified as ambiguous products. Median follow-up was higher at 631 days (IQR 365-729) in vaccinated individuals compared to 303 (IQR 92-656) in unvaccinated individuals.

Uptake with at least one dose declined with increasing age (**Figure 2; Supplementary Table S6;** 20.7% at 70 years vs 9.8% at 78 years; OR 0.41, 95% CI 0.35-0.48), albeit with an attenuation of this trend at 79 years (15.8% OR 0.75, 95% CI 0.67-0.83). Uptake was lower in women than men (17.1% vs 17.7%; OR 0.94, 95% CI 0.90-0.98). Uptake was higher in individuals with lower-risk immunosuppression, compared to higher-risk conditions (19.6% vs 14.3%; OR 1.48, 95% CI 1.40-1.56). Across immunosuppression-risk-level subcategories, uptake was 18.4% in Subcategory 1 (bone marrow compromising conditions), 19.1% in Subcategory 2 (solid organ transplant; OR 1.05, 95% CI 0.90-1.23), 12.4% in Subcategory 3 (recent cancer treatment; OR 0.61, 95% CI 0.56-0.67), 17.8% in Subcategory 4 (non-cancer immunosuppressive treatment; OR 0.93, 95% CI 0.86-1.01), and 22.1% in Subcategory 5 (immunodeficiency; OR 1.27, 95% CI 1.17-1.37), with longer follow-up observed in subcategories with higher uptake (**Supplementary Table S7**).

**Figure 2.**
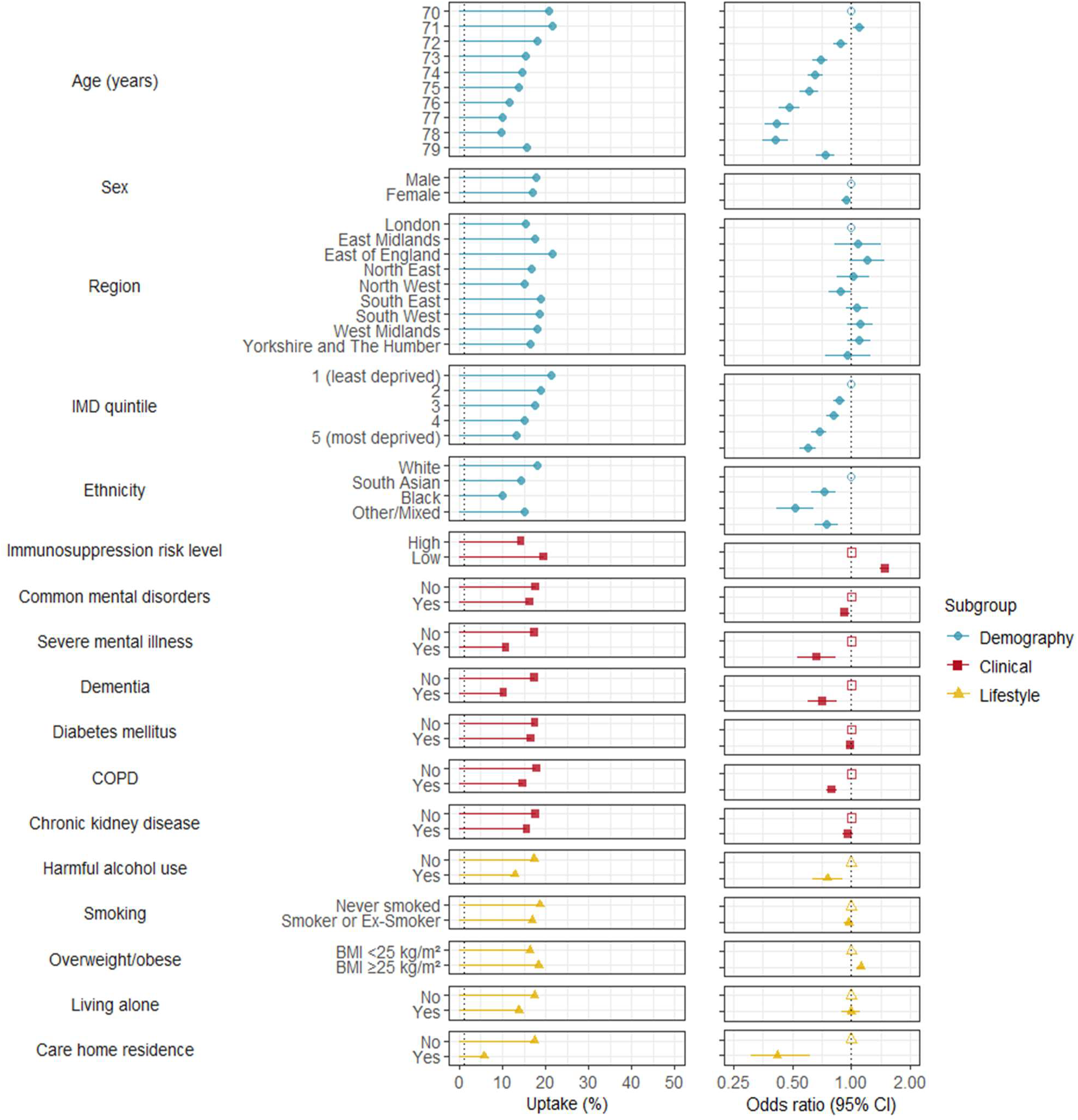
Association between uptake-related factors and receipt of at least one Shingrix^®^ vaccine in immunosuppressed adults. **Left panel:** Absolute coverage for individuals with and without the uptake-related factor of interest (%). **Right panel:** Odds ratios (95% confidence intervals) for each uptake-related factor. All odds ratios were adjusted for the following variables (i.e., main analysis model): age, sex, shingles infection in the last year, region, deprivation, ethnicity, immunosuppression risk level, common mental disorders, severe mental illness, dementia, diabetes, COPD, CKD, harmful alcohol use, smoking, overweight/obese. Analyses included 61,957 individuals with complete data. **Abbreviations:** IMD, index of multiple deprivation quintiles; CMD, common mental disorders; SMI, severe mental illness; DM, diabetes mellitus; COPD, chronic obstructive pulmonary disease; CKD, chronic kidney disease; BMI, Body Mass Index.

Uptake was lower: 1) among minority ethnic groups (18.0% in White individuals, 10.0% in Black individuals [OR 0.53, 95% CI 0.43-0.66], 14.2% in Asian individulas [OR 0.72, 95% CI 0.62-0.83], and 15.1% in individuals with mixed or other ethnicity [OR 0.75, 95% CI 0.65-0.86]); 2) with higher deprivation (13.0% in the most deprived vs 21.3% in least deprived IMD quintiles [OR 0.61, 95% 0.56-0.67]); 3) in individuals with records suggesting harmful alcohol use (12.9% vs 17.4% without [OR 0.74, 95% CI 0.62-0.89]); and 4) in people with severe mental illness (10.8% vs 17.4% without [OR 0.67, 95% CI 0.53-0.84]), dementia (10.3% vs 17.5% without [OR 0.71, 95% CI 0.60-0.84]), and COPD (14.8% vs 17.9% without [OR 0.79, 95% CI 0.75-0.84]). Care home residence uptake was also particularly low (5.8% vs 17.5% in those not in care homes [OR 0.44, 95% CI0.31-0.62]).

Associations for individual uptake-related factors were generally consistent across sequentially adjusted models (**Supplementary Table S6**). Results of sensitivity analyses were broadly similar to our main analysis (**Supplementary Tables S7, S8, S9**).

### Secondary analyses

#### Two-dose uptake

Among individuals who received the first dose and had at least 13 months of follow-up after first vaccination, 41.5% (2,307/5,557) completed the two-dose vaccine course between 7 weeks and 13 months. Among individuals with at least 13 months of follow up, the characteristics of those who received only one dose and those who received both were largely similar (**Supplementary Table S10**). The median interval between the first and second doses was 63 days (IQR: 57-77 days). Of the second doses administered, 2.3% (54/2,307) were received between 7 and 8 weeks after the first dose, 94.1% (2,171/2,307) were received within the guideline-recommended window of 8 weeks to 6 months, and 3.6% (82/2,307) were received more than 6 months after the initial dose (i.e., beyond the recommended interval).

#### Inadvertent Zostavax^®^

Only 1.0% (902/86,197) of individuals in our cohort of older adults with immunosuppression received Zostavax^®^. Eight-hundred and eighty seven first doses were Zostavax^®^, and 20 were administered as second vaccines (between 7 weeks and 13 months after the first).

#### Co-administration with the influenza vaccine

Among 9,888 individuals who received their first zoster vaccine between September and March (inclusive) of any year: 6.8% (n=676/9,888) received an influenza vaccine on the same day as their zoster vaccination; 2.4% (n=242/9,888) received it within 1-6 days either side; and 7.0% (n=695/9,888) received it within 7-14 days either side of Shingrix^®^ vaccination.

#### Ethnicity and deprivation

We explored whether any association between ethnicity and Shingrix^®^ uptake was driven by deprivation, by describing uptake in each ethnic group within strata of IMD quintiles. We found no evidence of interaction between ethnicity and IMD on Shingrix^®^ uptake (**Supplementary Table S11**; likelihood ratio test, p = 0.812).

## Discussion

### Summary

Our findings suggest that approximately 17% of eligible immunosuppressed adults received at least one Shingrix^®^ vaccination during the initial rollout (2021-2023), with 42% of those vaccinated going on to receive a second vaccine dose (in those who were followed up for at least 13 months after the first vaccine). We found that people were less likely to be vaccinated if they were older, more deprived, of minority ethnicity, had evidence suggesting harmful alcohol use, mental health conditions (including common mental disorders and severe mental illness), dementia, COPD, were considered at higher risk due to the reason for their immunosuppression, or were care home residents. Conversely, people were more likely to be vaccinated if they were overweight or obese.

### Findings in context

Our findings align with previous research highlighting disparities in the uptake of various vaccines across sociodemographic and clinical factors in the UK.(13) In a previous Zostavax^®^ uptake study in the UK general population (2013 to 2015), while overall uptake at 53% was higher than in our population of immunosuppressed individuals, the study demonstrated similar uptake-related factors to those found in our study (e.g., lower uptake in older people, more deprived individuals, those of minority ethnicity, and care home residents).

Similar disparities in vaccine uptake among immunosuppressed individuals to those observed in our study have been seen in other immunisation programmes, including influenza and COVID-19.(23,24) A 2022 paper of 3-dose COVID-19 vaccine uptake in people with kidney transplants saw lower uptake associated with minority ethnicity, higher social deprivation, and severe mental illness.(23) A recent study of the older population in the UK examined vaccine uptake rates for influenza, pneumococcus, and zoster between 1989 and 2020.(24) Zoster vaccine uptake was 53%, with lower uptake among minority ethnic groups and in more deprived populations, and higher uptake in individuals with more comorbidities. Individuals on immunosuppressive medication were less likely to receive a zoster vaccine, reflecting that the live Zostavax^®^ in use during the study period is contraindicated in those with immunosuppression.

The UK Health Security Agency (UKHSA) estimated first-dose Shingrix^®^ uptake at 19% in immunosuppressed individuals who turned 50 and over from September 2023 to August 2024 (as the UK shingles vaccination programme for immunosuppressed individuals expanded from September 2023 to include those aged 50 and over).(25) These updated UKHSA uptake estimates indicate the suboptimal uptake seen in our results has persisted following the expansion of the zoster vaccination programme to younger immunosuppressed individuals. Similar to our results in the immunosuppressed population, the UKHSA estimates indicated that for the general population turning 71 from April 2023 to March 2024, uptake of zoster vaccination as of 31 March 2024 varied markedly by ethnicity, with the highest uptake observed among those with White British ethnicity and lowest in Black or Black British-Caribbean ethnicity.

### Strengths and limitations

Our study has strengths, including using a large, nationally representative population, and comprehensive routinely-collected primary care data. Our detailed operationalisation of immunosuppression based on Green Book guidelines ensured our study population was clinically relevant. Our use of linked deprivation data enabled robust assessment of socioeconomic disparities.

Our study also had limitations. We may not have been able to reliably operationalise the Green Book definition of severe immunosuppression. For example, certain immunosuppressive treatments that are mainly prescribed in hospitals (e.g., biological agents), will not be completely captured in primary care data. However, our immunosuppression definition is based on code lists developed in conjunction with UKHSA to help identify individuals using primary care electronic health records who broadly fit eligibility criteria for specific vaccine programmes.(26) Therefore, we expect that primary care practices will have used a broadly comparable approach to identify individuals potentially eligible for Shingrix^®^. So, while our immunosuppression definition may not identify all immunosuppressed individuals, it should have captured those identified by the primary care clinicians who are administering vaccinations, and consequently reflect disparities in uptake.

Our assignment of immunosuppression-risk-level subcategories reflects recent work assessing risk hierarchy for COVID-19 outcomes (11,12) that may not accurately capture shingles risk (e.g., given the prominent role of T cells in preventing zoster reactivation (27)). Although uptake was lower in individuals with higher-risk conditions, this may have been driven by lower uptake in Subcategory 3 (recent cancer treatment) that was associated with a shorter follow-up period than other immunosuppression-risk-level subcategories. We defined active cancer based on a coded diagnosis in primary care in the past 12 months, which we deemed to be a sensitive proxy for Green Book eligibility criteria. However, our definition may not align directly with the target population invited for vaccination if based specifically on records of cancer treatments (we may have underestimated the target population if individuals receiving cancer diagnoses and treatments given in secondary care settings did not have timely recording of their diagnosis in primary care records). Follow-up time was also shorter in Subcategory 3 than other immunosuppression-risk-level subcategories, which may contribute to the lower uptake observed in this group. In addition, some of the newer targeted immunosuppressive therapies included in Subcategory 4 (Non-cancer immunosuppressive treatment) and conditions included in Subcategory 5 (Immunodeficiency) may have substantial effects on immune function, and therefore may not necessarily reflect lower risk. Overall, our findings on immunosuppression risk level should be interpreted with caution given uncertainty over how best to operationalise and stratify Green Book risk groups in electronic health records. Efforts to develop and validate expert consensus definitions of immunosuppression phenotypes are ongoing.(28)

Incomplete data on ethnicity, deprivation, overweight/obese, and smoking status may have introduced selection bias (as we used a complete case analysis). However, results from earlier models that did not adjust for these factors were broadly similar to main analysis results. Care home residence and living alone were defined based on morbidity coding and coded as negative if no code was recorded, which is likely to underestimate the true number living alone and in care homes.

### Possible explanations and implications for policy and practice

Overall uptake in those with immunosuppression remains low at 17%. This low uptake is consistent across all demographic and clinical characteristics examined in this study, raising concerns about the effectiveness of communication efforts directed at people with immunosuppression regarding their eligibility for vaccination, as well as constraints in vaccine delivery capacity. In England, individuals identified as immunosuppressed are eligible for vaccination and, in principle, should receive personalised invitations to book appointments via letters, text messages, or direct contact from their general practice.(29) However, in practice, the identification and invitation processes may be inconsistent, as they rely on accurate coding of immunosuppression in primary care records and local implementation by practices and vaccination services.

Low zoster vaccine uptake should be considered within the broader context of the COVID-19 pandemic. The zoster vaccine guidelines changed in September 2021, just before the emergence of the COVID-19 Omicron variant, which led to a case surge during the winter of 2021/22. During this period, much of the public health focus was on rolling out COVID-19 booster doses, which may have overshadowed efforts to promote zoster vaccination. Additionally, health system capacity was strained due to high COVID-19 infection and hospitalisation, which could have limited the ability of healthcare providers to focus on other preventative healthcare. It has been suggested that repeated vaccination campaigns within a short time frame can lead to declining motivation to get vaccinated.(30) By late 2021, many individuals had already received multiple COVID-19 vaccine doses, which may have resulted in fewer people seeking additional vaccinations.(30)

Our findings have important implications for public health policy and clinical practice. Firstly, we identified decreasing uptake with increasing age. Lower uptake with increasing age highlights potential for increasing uptake in catch-up cohorts, as well as those starting to become eligible. The lower uptake of Shingrix^®^ among minority ethnic groups and more deprived individuals suggests a need for tailored outreach strategies to improve access and acceptance in underserved populations. For example, enhanced primary care engagement may address barriers faced by minority ethnic groups. Lower uptake in those with CMD, SMI and dementia may suggest a need for tailored strategies to support vaccine uptake in these vulnerable individuals. Further research should explore the underlying reasons for disparities in Shingrix^®^ vaccination, particularly among already underserved populations. Qualitative studies involving people with immunosuppression and their healthcare providers could provide valuable insights into barriers and facilitators of uptake in this vulnerable group. Additionally, longitudinal analyses following the expanded vaccination programme in 2023 could assess its impact on reducing disparities and improving coverage among high-risk groups.

When Shingrix^®^ was first introduced in 2021, the Green Book guidelines indicated that the vaccine could be co-administered with inactivated influenza vaccine, but recommended against routinely offering the vaccine at the time as the adjuvanted influenza vaccine due to the absence of safety data.(7) These guidelines have been updated in light of reassuring data for co-administration of Shingrix^®^ with adjuvanted influenza vaccine.(31) Co-administration of Shingrix^®^ with the influenza vaccine presents an opportunity to improve coverage through opportunistic vaccination, and practices could further integrate vaccine delivery to optimise patient visits for routine care.

## Conclusion

Our study highlights disparities in Shingrix^®^ vaccination uptake among immunosuppressed individuals, with low overall coverage and particularly reduced uptake in older people, those living in more deprived areas, individuals from minority ethnic groups, and people with dementia or severe mental illness. While many factors were associated with reduced uptake in relative terms, only a subset showed large absolute differences, most notably ethnicity, deprivation, those living with dementia or severe mental illness, and care home residence underscoring areas of substantial inequality in vaccine delivery. Our findings are consistent with patterns seen for other vaccines and reinforce the need for targeted outreach and tailored strategies to improve access in underserved groups. Integration of Shingrix^®^ with other routine vaccinations, such as the influenza vaccine, may offer an opportunity for improving coverage.

## Data Availability

The study uses data from the Clinical Practice Research Datalink (CPRD). CPRD does not allow the sharing of patient-level data. The data specification for the CPRD dataset build used in this paper is available at: https://www.cprd.com/doi/cprd-gold-june-2024-dataset. Analysis code and code lists are shared in our online Github repository at doi:10.5281/zenodo.15351553.

https://doi.org/10.5281/zenodo.15351553

## Acknowledgements

This study is based in part on data from the Clinical Practice Research Datalink obtained under licence from the UK Medicines and Healthcare products Regulatory Agency. The data is provided by patients and collected by the NHS as part of their care and support. The interpretation and conclusions contained in this study are those of the authors alone.

## Contributors

KM, EP, ID, and NA contributed to Conceptualisation. EB, EP, KM, and AS contributed to Data Curation. EB and EP were responsible for Formal Analysis. EB and EP conducted the Investigation. Methodology was developed and designed by all authors. EB, KM, and EP contributed to Project Administration. AS, KM, and ML contributed to Resources. EB, KM, AS, and EP worked on Software, including programming, developing code, and implementing computational tools. KM and EP provided Supervision. EB and EP supported Validation, helping to verify and ensure the reproducibility of results and research outputs. Visualisation was carried out by EB and EP, who created and presented data visualizations for the published work. EB, EP, and KM contributed to Writing - Original Draft. All authors participated in Writing - Reviewing and Editing, providing critical review, commentary, and revisions at various stages of the publication process. EB is the guarantor. The corresponding author (EB) attests that all listed authors meet authorship criteria and that no others meeting the criteria have been omitted.

## Patient consent for publication

Not required.

## Ethics approval

The study was approved by the Independent Scientific Advisory Panel of the CPRD (protocol reference 24_003902) and the London School of Hygiene and Tropical Medicine Ethics Committee (LSHTM reference 30861). The study protocol was made available to reviewers

## Data availability and sharing

The study uses data from the Clinical Practice Research Datalink (CPRD). CPRD does not allow the sharing of patient-level data. The data specification for the CPRD dataset build used in this paper is available at: https://www.cprd.com/doi/cprd-gold-june-2024-dataset. Analysis code and code lists are shared in our online Github repository at doi:10.5281/zenodo.15351553.(15)

## Open access

This is an open access article distributed in accordance with the Creative Commons Attribution 4.0 Unported (CC BY 4.0) license, which permits others to copy, redistribute, remix, transform and build upon this work for any purpose, provided the original work is properly cited, a link to the licence is given, and indication of whether changes were made. See: https://creativecommons.org/licenses/by/4.0/.

