## Supplemental information for "Disparities in uptake of Shingrix^®^ vaccine in immunosuppressed individuals in England: a population-based cohort study"

#### Supplementary Methods

##### Immunosuppression definitions

We used the University of Nottingham Primary Care Information Services (PRIMIS) data specification for immunosuppression (commissioned by UKHSA) to develop our immunosuppression definition code lists.(1–3) PRIMIS have worked with the UKHSA to develop code lists and logical business rules (pseudocode) to help identify individuals using primary care electronic health records who broadly fit Green Book defined eligibility criteria for specific vaccine programmes. However, the PRIMIS data specification for Shingrix® is only intended to identify people potentially eligible for Shingrix® (based on the Green Book Shingrix® eligibility criteria for immunosuppression). It is then a clinical decision regarding whether identified individuals meet the criteria.(4–7) Therefore, where necessary, we refined algorithms, to capture the Green Book criteria more closely. We reviewed all PRIMIS code lists against other existing code lists developed based on Green Book definitions of immunosuppression for receipt of flu and COVID vaccinations (where necessary mapping existing Read-code lists to SNOMED codes). While we appreciate that there may be differences between the definitions of immunosuppression used as indications for other vaccine immunisation programmes (i.e., influenza, COVID-19), we felt that a thorough review of existing code lists allowed us to identify relevant codes to capture the eligible immunosuppressed population more robustly.

We acknowledge that we may not have been able to fully operationalise the Green Book criteria for the eligible immunosuppressed population using CPRD as some data was not recorded. For example, we were not able to fully capture receipt of targeted biologics in primary care data as these are mainly delivered through secondary care, and linked secondary care prescribing data are often not available.

Operationalising the Green Book guidance for duration of immunosuppression using electronic health records (EHRs) is not straightforward. In EHRs, records indicating the resolution of a diagnosis or the end of a treatment are often not available. We were also not able to reliably capture test results and prescriptions undertaken in secondary care. Therefore, we applied pre-defined time periods for immunosuppression duration (based on those used in previous studies that were developed based on the implied durations of immunosuppression presented in the Green Book immunosuppression criteria) with additional time added where necessary to allow for the lack of resolution recording in primary care records.(4–6) Consequently, in some instances, the Green-book defined periods of immunosuppression may not completely align with our EHR-based operationalisation of them due to the limitations of routinely recorded primary care health record data.

Our condition-specific durations of immunosuppression were (further details justifying choice of immunosuppression duration are available in **Supplementary Table S1**):

1. Primary or acquired immunodeficiency:
    - Haematological malignancies, including leukaemia, lymphoma, and other lymphoproliferative disorders, were considered immunosuppressed for 24 months following a record.
    - HIV/AIDS: permanently considered immunosuppressed (as we cannot reliably establish current CD4 cell count in primary care data).
    - Cellular immune deficiencies: aplastic anaemia, considered immunosuppressed for 24 months following a relevant record; congenital cellular immune deficiencies, considered permanently immunosuppressed; and those with other/unspecified cellular immune deficiencies, considered immunosuppressed for 3 months.
2. Stem cell transplant: permanently considered immunosuppressed (as we could not reliably establish whether there was ongoing immunosuppression or graft versus host disease).
3. Immunosuppressive or immune-modulating therapy (chemotherapy, radiotherapy, targeted immunotherapy):
    - Chemotherapy or radiotherapy: considered immunosuppressed for 1 year from record.
    - Solid organ transplants: considered to be immunosuppressed permanently following transplant, as almost all organ recipients remain on anti-rejection medications for life.

- Targeted therapy for autoimmune disease (biologics): 12 months following each prescription.

4. Other immunosuppressive medications for immune-mediated inflammatory disease (excluding cancer-related immunosuppressive medications, biologics and small molecules, and low-dose corticosteroids):
   - Moderate-to-high dose corticosteroids (≥20mg prednisolone equivalent dose per day) for more than 10 days (do not need to be consecutive) in the previous month: considered immunosuppressed for 3 months last corticosteroid dose.
   - Long-term moderate dose corticosteroids (equivalent to ≥10mg prednisolone per day for more than 4 weeks, days do not need to be consecutive) in the previous 3 months: considered immunosuppressed for 3 months last corticosteroid dose.
   - Non-biological immune modulating drugs (e.g., methotrexate, azathioprine, 6-mercaptopurine) were considered immunosuppressed for 3 months after the last dose.
5. Short-term high-dose corticosteroids (≥40mg prednisolone equivalent dose per day for more than 7 days [do not need to be consecutive] for any reason in the previous month: considered immunosuppressed for 3 months from last dose.

We defined all corticosteroid prescribing definitions (i.e., moderate-to-high dose, long-term moderate, and short-term high-dose) with a duration of 3-months from end of last dose, as it can be complex to establish corticosteroid dosing using primary care EHR data. Corticosteroids are often prescribed with reducing doses, to allow an individual to taper their steroid dose over time to avoid some of the problems with suddenly stopping a high dose (e.g., 40mg daily for a week, 30mg daily the next week, 20mg the next, 10mg the next, and a final week at 5mg per day). In this situation, an individual may be issued with multiple different strength tablets of a corticosteroid on the same day, with clear instructions for how to taper their dose over time that may not be captured in the EHRs. Consequently, it can sometimes be difficult to establish whether a clinician intended multiple different strength tablets prescribed on the same day to be taken starting on that day, or as part of a tapered regimen. Therefore, we took the longest period associated with corticosteroid prescribing in the Green Book criteria and applied it to our definition of the end of immunosuppression.

##### Harmful alcohol use

We defined harmful alcohol use based on records for any primary care morbidity codes suggesting harmful or heavy alcohol use (including harmful alcohol use codes and codes related to physical/psychological harm related to alcohol use), or any prescription for drugs used to maintain abstinence recorded before study entry (products included the following generic/brand named drugs: acamprosate, disulfiram, nalmefene, Campral, Antabuse, Esperal, Selincro). Morbidity code lists were based on those used in a previous publication, excluding family history.(8) In our main analysis, our definitions were based on records before cohort entry.

##### Smoking

We defined smoking status based on primary care morbidity coding. We categorised smoking status as: non-smoker, smoker or ex-smoker, or missing. Smoking status was defined based on records from the 10 years prior to study entry. Individuals coded as “non-smoker” with no entries indicating smoking or ex-smoking status were labeled as non-smokers. If there was no smoking-related coding within the last 10 years, the individual was recorded as having missing smoking data.

##### Overweight/obese

We considered individuals with a body mass index (BMI) of 25 $kg/m^2$ or more, recorded on or before cohort entry, as overweight or obese. This was calculated using the height and weight measures, recorded before cohort entry date. If there were multiple height and weight measures recorded on different days, we used those recorded closest to cohort entry date. We excluded:

1. Values recorded in implausible units, for example if values were not consistent with standard measurements in kilograms or meters.
2. Extreme values:

- Weight: retained only if between 20kg and 400kg
- Height: retained only if between 1.2m and 3.0m

If multiple discordant measurements were recorded on the same day, all measurements for that day were discarded to reduce the likelihood of erroneous data entry if:

- The difference in height exceeded 0.5m, or
- The difference in weight exceeded 5kg, or

Otherwise, if multiple measures recorded on the same day did not meet the criteria above, we used the mean BMI calculated from all measures available on that date.

**Chronic Kidney Disease (CKD)**

We used serum creatinine values alongside age and sex to calculate estimated glomerular filtration rate (eGFR) using the CKD-EPI equation without ethnicity.(9,10) Individuals were classified as having CKD if their most recent eGFR measure before cohort entry was 60mL/min/1.73 $m^2$ or under.

Supplementary Table S1: Detailed immunosuppression definitions based on Green Book criteria for severe immunosuppression.

| Hierarchical risk category | Hierarchical level | Green Book defined immunosuppression category | Condition/drug | Green Book criterion (verbatim)* | Definition operationalised for electronic health records algorithm | | |
|---|---|---|---|---|---|---|---|
| | | | | | Dose criteria if applicable** | Remains immunosuppressed until | Notes/justification |
| 1. Bone marrow compromising conditions | High | Primary or acquired immunodeficiency | Haematological malignancies, including leukaemia, lymphoma, and other lymphoproliferative disorders | Acute and chronic leukaemia, and clinically aggressive lymphoma (including Hodgkin's lymphoma) who are less than 12 months since achieving cure | N/A | 24 months after a record | An additional 12 months added, as we were unlikely to see a code for 'cure'. |
| | | | | Individuals under follow up for chronic lymphoproliferative disorder including haematological malignancies such as indolent lymphoma, chronic lymphoid leukaemia, myeloma, Waldenstrom's macroglobulinemia and other plasma cell dyscrasias (N.B: this list not exhaustive) | N/A | 24 months after a record | Followed for 24 months from a record as we were unlikely to be able to establish whether someone is 'under follow up' from the data. |
| | | | Stem cell transplant | Those who have received an allogeneic (cells from a donor) or an autologous (using their own cells) stem cell transplant in the previous 24 months | N/A | Permanent | While the Green Book criterion mentions 24 months for stem cell transplant without continuing immunosuppression or graft versus host disease (GVHD), we could not reliably establish continuing immunosuppression/GVHD. So, we conservatively set individuals with stem cell transplant to be immunosuppressed permanently. |
| | | | | Those who have received a stem cell transplant more than 24 months ago but have ongoing immunosuppression or graft versus host disease (GVHD) Individuals | | | |

| | | | | | | | |
|---|---|---|---|---|---|---|---|
| 2. Solid organ transplant | High | Immunosuppressive or immune-modulating therapy (chemotherapy, radiotherapy, targeted immunotherapy | Solid organ transplant | Those who are receiving or have received in the previous 6 months immunosuppressive therapy for a solid organ transplant | N/A | Permanent | Pragmatically considered to be immunosuppressed permanently following transplant, as almost all organ recipients will remain on anti-rejection medications for life. |
| 3. Recent cancer treatment | High | Immunosuppressive or immune-modulating therapy (chemotherapy, radiotherapy, targeted immunotherapy | Recent cancer diagnosis | Those who received in the past 6 months immunosuppressive chemotherapy or radiotherapy for any indication | N/A | 12 months | Green Book criteria specify that individuals remain immunosuppressed for 6 months after immunotherapy. However, many individuals would start this treatment in secondary care and therefore primary care records are likely to miss the true start of treatment. We therefore cautiously assumed that individuals remained immunosuppressed for up to 12 months from initial cancer diagnosis. |
| 4. Non-cancer immunosuppressive treatment | Low | Other immunosuppressive medications for immune-mediated inflammatory disease (excluding cancer-related immunosuppressive medications, biologics, and low-dose corticosteroids): | Moderate-to-high dose corticosteroids | Moderate to high dose corticosteroids (equivalent ≥20mg prednisolone per day) for more than 10 days in the previous month | ≥20mg prednisolone equivalent dose more than 10 days in a month | 3 months after last dose | While the Green Book criterion specifies prescribing over a month, we conservatively used 3 months after the last dose, given the complexities of establishing steroid dosing in electronic health records. |
| | | | Long-term moderate-dose corticosteroids | Long term moderate dose corticosteroids (equivalent to ≥10mg prednisolone per day for more than 4 weeks) in the previous 3 months | ≥10mg prednisolone equivalent dose more than 4 weeks in 3 months | 3 months after last dose | |

| | | | | | | | |
|---|---|---|---|---|---|---|---|
| | | | Non-biological immune modulating drugs | Any non-biological oral immune modulating drugs e.g., methotrexate >20mg per week (oral and subcutaneous), azathioprine >3.0mg/kg/day; 6-mercaptopurine >1.5mg/kg/day, mycophenolate >1g/day) in the previous 3 months | N/A | 3 months after last dose | While Green Book criteria indicate specific minimum doses for some non-biological immune modulating drugs to be defined as leading to severe immunosuppression, we operationalised our definition based on any dose. We acknowledge that this approach of defining immunosuppression based on any dose of immune modulating drugs will include some individuals on lower doses whose immune systems may not be as strongly suppressed as those on higher doses. However, it would have been difficult to robustly operationalise a weight-based dose definition in primary care records (as weights are not routinely recorded in primary care data, and when multiple weight measures are recorded it would be difficult to decide which measure to use). Additionally, in clinical practice, lower doses of these drugs tend to be used in more frail individuals who are likely to have less resilient immune systems anyway.<br><br>We included the following drugs in our definition: methotrexate, azathioprine, mercaptopurine, mycophenolate, leflunomide, and ciclosporin. |

| | | | | | | | |
|---|---|---|---|---|---|---|---|
| | | | Combination therapies | Certain combination therapies at individual doses lower than stated above, including those on ≥7.5mg prednisolone per day in combination with other immunosuppressants (other than hydroxychloroquine or sulfasalazine) and those receiving methotrexate (any dose) with leflunomide in the previous 3 months | N/A | Not included in our immunosuppression algorithm. | We defined individuals as immunosuppressed based on any dose of non-biological immune modulating drug (see row above). Consequently, individuals meeting the Green Book severe immunosuppression criteria based on combination prescribing, were included in how we operationalised the non-biological immune modulating drugs criterion (in the row above). Our definition therefore does not explicitly operationalise this combination-prescribing Green Book criterion for severe immunosuppression. |

| | | | | | | | |
|---|---|---|---|---|---|---|---|
| | | Immunosuppressive or immune-modulating therapy (chemotherapy, radiotherapy, targeted immunotherapy | Targeted immunotherapy (i.e., biologics or small molecule drugs)*** | Those who are receiving or have received in the previous 3 months targeted therapy for autoimmune disease, such as JAK inhibitors or biologic immune modulators including B-cell targeted therapies (including rituximab but for which a 6 month period should be considered immunosuppressive), monoclonal tumor necrosis factor inhibitors (TNFi), T-cell co-stimulation modulators, soluble TNF receptors, interleukin (IL)-6 receptor inhibitors., IL-17 inhibitors, IL 12/23 inhibitors, IL 23 inhibitors (N.B: this list is not exhaustive) | N/A | 12 months following a prescription | These drugs were unlikely to be well captured in primary care records, as most were administered in secondary care. However, inclusion of those individuals identified based on primary care recording ensured that we included as many people with immunosuppression in our study population as possible.<br><br>While Green Book criteria specify immunosuppression in 3 months following targeted immunotherapy, we pragmatically considered individuals to be immunosuppressed for 12 months from a record for one of these drugs due to the poor capture of the drugs in primary care records (and rituximab is considered to lead to immunosuppression for 12 months from receipt). |
| | | Short-term of high-dose corticosteroids | | Individuals who have received a short course of high dose steroids (equivalent >40mg prednisolone per day for more than a week) for any reason in the previous month. | >40mg prednisolone per day for more than a week | 3 months after last dose | While the Green Book criterion specifies prescribing over a month, we conservatively followed for 3 months after the last dose, given the complexities of establishing steroid dosing in electronic health records. |
| 5. Immunodeficiency | Low | Primary or acquired immunodeficiency | HIV/AIDS | immunosuppression due to HIV/AIDS with a current CD4 count of below 200 cells/µl. | N/A | Permanent | We cannot reliably establish CD4 count in primary care, as it is likely most monitoring and testing will happen in secondary care. Therefore, we regarded anyone with a record of HIV/AIDS as being immunosuppressed. |

| | | | | | | | |
|---|---|---|---|---|---|---|---|
| | | Primary or acquired immunodeficiency | Cellular immune deficiencies | primary or acquired cellular and combined immune deficiencies – those with lymphopaenia (<1,000 lymphocytes/ul) or with a functional lymphocyte disorder | N/A | Permanent | Periods of immunosuppression based on clinical reasoning. |

*As defined in Green Book, Shingrix® Chapter (11)

** Where specific daily doses were required to be more than a set number of days/weeks in a specified period, the days do not have to be consecutive.

*** Targeted immunotherapy include the following drugs: abatacept, adalimumab, aflibercept, alemtuzumab, anakinra, apremilast, thymoglobulin, baricitinib, basiliximab, belatacept, bevacizumab, bortezomib, brentuximab, canakinumab, cetuximab, certolizumab, daclizumab, dasatinib, eculizumab, etanercept, everolimus, fingolimod, golimumab, idelalisib, imatinib, infliximab, ipilimumab, natalizumam, nilotinib, nivolumab, obinutuzumab, ofatumumab, panitumumab, pembrolizumab, pertuzumab, rituximab, secukinumab, temsirolimus, tocilizumab, tofacitinib, trastuzumab, ustekinumab, vedolizumab, abrocitinib, upadacitinib, dupilumab, tralokinumab, gudelkumab, brodalumab, ixekizumab, bimekizumab, rizankizumab.

#### Supplementary Table S2: Operationalisation of **non-biological immune-modulating drugs** definitions for immunosuppression, including specific coding choices.

To operationalise our non-biological immune-modulating drugs definition of immunosuppression, we needed to identify the dose prescribed each day (daily dose strength) and duration of steroid prescriptions. We used data from:

1. **Prescription** records for each individual containing duration and quantity (but lacking in more exact data). We termed these **'base'** records.
2. The **common prescription linkage data** (can be linked to base prescription records), which contain daily dose, dose duration (regarded as a more accurate record of dose duration than that derived from prescription data) and description of prescription. We termed this **'linkage'** data. Only some records had common linkage.

| Step | Methodology |
|---|---|
| 1. Prescription duration | We defined the duration of each prescription as follows:<br>● The linkage file was only used when the daily dose was specified in a recognisable unit (e.g., mg, ml, g), otherwise we used duration from the base prescriptions.<br>● Linkage records containing terms like "needed" or "required" were removed, as this would indicate drugs taken as needed to resolve symptoms.<br>● If the daily dose was provided in dosage units (e.g., ml, mg, mcg) rather than discrete quantities, it was set to 0, meaning the calculation would rely on quantity and duration from the CPRD base prescription data instead.<br>● If available, we used the dose duration provided in the common dosage linkage file.<br>● If duration was not available from the linked data, it was calculated from the base prescription records as:<br>Duration='Quantity provided by CPRD prescription records'/'Daily dose from common dosage'<br>● If quantity was under 1 dose (but not 0), duration was set to 1 day.<br>● If no duration was assigned at this point, CPRD duration from the base prescription records (not the linkage file) was used.<br>● If duration was still missing, it was set to the median/mode duration of 28 days.<br>● If duration exceeded 365 days, it was capped at 28 days |
| 2. Handling of injections | Daily dosage was calculated as the number of units of a drug prescribed per day, multiplied by the substance strength for one unit of the drug (provided by CPRD in the base prescription records).<br>Daily dose was derived as follows:<br>● If a daily dose was explicitly provided in the linkage, it was used.<br>● If missing, it was calculated as:<br>Daily dose='Quantity provided by CPRD base prescription records'/'Derived duration' |
| 3. Defining immunosuppression period | An individual was considered immunosuppressed starting from the first day of prescription to 3 months after their last prescription. |

#### Supplementary Table S3: Operationalisation of **steroid definitions** for immunosuppression, including specific coding choices.

In order to operationalise our corticosteroid-based definition of immunosuppression, we needed to identify the dose prescribed each day (daily dose strength) and duration of steroid prescriptions, as well as accounting for any multiple prescriptions on the same day and overlapping prescriptions.

We used data from:

1. **Prescription** records for each individual containing duration and quantity (but lacking in more exact data). We termed these **'base'** records.
2. The **common prescription linkage data** (can be linked to base prescription records), which contain daily dose, dose duration (regarded as a more accurate record of dose duration than that derived from prescription data) and description of prescription. We termed this **'linkage'** data. Only some records had common linkage.

| Step | Methodology |
|---|---|
| 1. Systemic steroids | We only included systemic steroids. We therefore excluded any steroids administered through routes that would be unlikely to have a systemic effect, e.g., inhalation, topical application, or ophthalmic preparations (eye drops). |
| 2. Excluding injectable prescriptions | Prescriptions identified as injections were removed because their prescription duration and immunosuppressive effect (for example, as some of these injections may have been administered into joints) could not be reliably established. |
| 3. Prescription duration | The Green Book definition of immunosuppression related to steroid prescribing requires specific daily doses over a defined period. We therefore defined the duration of each prescription as follows:<br>● The linkage was only used when the daily dose was specified in a recognisable unit (e.g., mg, ml, g), otherwise the base duration was used.<br>● Linkage records with a description containing terms like "needed" or "required" were removed, as this would indicate drugs taken as needed to resolve symptoms.<br>● We used linkage data duration if this was available.<br>● If linkage data duration was missing, we calculated duration as:<br>Duration = 'Quantity provided by base prescription records'/ 'Daily dose from dosage linkage'<br>● If duration was still missing, duration was set to CPRD's base duration record (i.e. not the linkage).<br>● If no duration was found in the base records, the median duration of all prescriptions included in the data (28 days) was used.<br>● If the calculated duration exceeded 365 days, it was capped at 28 days (Department of Health recommend that the length of repeat prescription supply should not routinely be more than 28 days).<br>● If the quantity was under 1 unit, duration was set to 1 day.<br>● This provided us with a derived duration for each record. |
| 4. Daily dose | Daily dosage was calculated as the number of units of a drug prescribed per day, multiplied by the substance strength for one unit of the drug (provided by base prescription records).<br>Daily dose was derived as follows:<br>● If a daily dose was explicitly provided in the linkage data, it was used.<br>● If missing, it was calculated as:<br>Daily dose='Quantity provided by base prescription records'/'Previously derived duration' |
| 5. Prednisolone equivalent dose | ● We calculated daily dose, by multiplying prednisolone equivalent dose (See (12) or doi:10.5281/zenodo.15351553) for specific steroids prescribed by the substance strength prescribed, and then by the units per day from Step 4.<br>● To avoid including implausible dosages, we removed prescriptions for more than 100mg prednisone per day. |

| | |
|---|---|
| 6. Multiple prescriptions on the same day | • If two identical prescriptions were recorded for the same individual on the same day, they were assumed to be taken consecutively, so their durations were added.<br>• If two different doses were prescribed on the same day, the prednisolone equivalent daily doses were added, and duration was set to the maximum of the two records.<br>• If prescriptions overlapped, later prescriptions overrode earlier ones, with the new prednisolone equivalent daily doses starting at the beginning of the later record and continuing until its end date.<br>• We ensured that the total prescription duration for steroids prescribed on the same day did not exceed 365 days (as this would be unlikely to be plausible). In the situation where a calculated prescription duration exceeded 365 days, we set it to 365 as a maximum. |
| 7. Start and end dates | Periods considered to be immunosuppressed based on corticosteroid prescribing were defined for each individual for each type of the three corticosteroid prescribing patterns. We determined the start and end date when an individual was considered to be immunosuppressed based on corticosteroid prescribing (see **Supplementary Table S1**). |

#### Supplementary Table S4: Summary of hierarchical modelling structure (where 'x' is the potential uptake-related factor under investigation)

We adjusted sequentially based on a hierarchical conceptual framework developed for each specific uptake related factor of interest. Existing conceptual frameworks were adapted to describe the presumed hierarchical relationships between distal and proximal factors and vaccine uptake.

| Model | Justification |
|---|---|
| **Model 1:** Age + Sex (+ x), where x represents each uptake-related factor | Sex and age were considered potential determinants of zoster vaccine uptake and *a priori* confounders of sociodemographic, clinical and lifestyle uptake-related factors, and were therefore included in the first level of the hierarchy. |
| **Model 2:** Model 1 + recent zoster infection (+ x) | We then adjusted for recent shingles as a potential factor affecting the choice of whether, or not, to vaccinate. Guidelines recommend that individuals with shingles should wait until their symptoms have resolved before receiving shingles immunisation (Greenbook, Chapter 28a (up to 31 August 2023)) However, since shingles will likely offer a natural boosting effect, the benefit of offering a zoster vaccine immediately following recovery is unclear. Consequently, we adjusted for recent shingles in to account for it affecting the choice to vaccinate or not (e.g., because a clinician does not think it is worthwhile due to the natural boosting effect of shingles, or because an individual was offered vaccination while still experiencing the rash of shingles) and the potential for previous shingles to be related to the uptake-related factors investigated. . |
| **Model 3**: Model 2 + region + deprivation + ethnicity (+x) | We then adjusted for region, deprivation and ethnicity as we felt these were distal determinants that would affect, directly or indirectly, all other uptake-related factors. |
| **Model 4:** Model 3 + immunosuppression risk level (+ x) | We then adjusted for clinical factors that we expected to be impacted by sociodemographic factors and to potentially influence lifestyle factors. We added immunosuppression risk level (i.e.,whether someone was considered to be at high- or low-risk of experiencing severe shingles) first given that clinicians/individuals may be more likely to recommend/ask for a shingles vaccination if they were felt to be at greater risk (defined based on Green Book criteria, applying findings from work on Coronavirus disease 2019-related risk)[Leston et al]. |
| **Model 5**: Model 4 + CMD + SMI + dementia + DM + COPD + CKD (+ x) | We then added clinical factors other than immunosuppression. |
| **Model 6**: Model 5 + harmful alcohol use + smoking + overweight/obese (+ x) | We then adjusted for lifestyle factors as a proximal determinant of uptake, anticipating potential collinearity with clinical factors. |
| **Model 7**: Model 6 + living alone + care-home residence | Finally, we adjusted for living alone and care-home residence status. We expected to be under-recorded in primary care records, so only adjusted for them in our final model to avoid introducing bias. |

#### Supplementary Table S5: Summary of sensitivity analysis

| Sensitivity | Justification |
|---|---|
| 1. Static cohort | In our main analysis individuals entered the study population on a rolling basis from the latest of when they meet the age criteria (i.e., 70-79) and our definition of immunosuppression. This dynamic approach to cohort entry allowed us to: 1) better capture individuals who may be offered vaccination opportunistically due to a change in their clinical status; and 2) include in our denominator individuals who were likely to be at great risk of zoster due to new diagnosis or treatment. However, our approach of dynamically defining our study population throughout the year, may not reflect how the shingles vaccination programme was operationalised by some primary care practices. Some practices may have implemented the shingles vaccination programme on an annual basis by identifying all eligible individuals and inviting them for vaccination at one time in the year. Our approach of including individuals who meet the immunosuppression definition after the start of the vaccination year may therefore unfairly inflate the denominator population for those practices implementing the vaccination programme through a single annual call up, making coverage look less complete than it is. Consequently, in this sensitivity analysis, we defined eligibility for the entire cohort on 1st September 2021 based on birth year (i.e., age 70-79 in 2021, born 1942-1951) and immunosuppression status as of 1st September 2021. In this sensitivity analysis, the cohort was static, with no additional individuals joining if they met our immunosuppression definition during the study period. . |
| 2. Time updated covariates | In our main analyses, all covariates were defined based on status recorded on or before start of follow up (i.e., when an individual became eligible for Shingrix® based on reaching 70 years of age or becoming immunosuppressed). To test the impact of this approach, we repeated the analysis based on time-updated covariate status recorded at any time during follow up.<br>We time updated status of: immunosuppression risk category, common mental disorders, severe mental illness, dementia, diabetes mellitus, chronic obstructive pulmonary disease (COPD), chronic kidney disease (CKD), harmful alcohol use, smoking history, body mass index (kg/m2), and living alone and care home residence. |
| 3. Restricted to at least 13 months follow-up | In our main analysis we did not apply any requirement for individuals to have a defined period of follow up. As individuals with limited follow up may not have sufficient time to receive a vaccination (therefore inflating the denominator population), in this sensitivity analysis, we restricted to individuals with at least 13 months follow up available after cohort entry, to allow sufficient time for individuals to have their vaccinations. |
| 4. Restricted to age 70 only | We restricted the analysis to those who were 70 at entry to the study, in order to study the age-defined individuals who were specifically targeted for Shingrix(R) vaccination at the time of the study. |
| 5. Sixteen categories of ethnicity | For sufficient statistical power in the main analysis, we considered four different ethnicity categories. In a sensitivity analysis, we considered sixteen ethnicity categories to highlight potential disparities in uptake in more narrowly defined ethnic groups. |

| | |
|---|---|
| 6. Five immunosuppression risk categories | In our main analysis we used two immunosuppression risk-level categories : higher (bone marrow compromised, solid organ transplant) and lower risk (recently treated cancer, non-cancer immunosuppressives, immunodeficiency). In this sensitivity analysis we investigated whether results varied using five levels of immunosuppression: 1) bone marrow compromised; 2) solid organ transplant; 3) recently treated cancer; 4) non-cancer related immunosuppressives; and 5) immunodeficiency (primary and acquired). |

#### Supplementary Table S6: Odds ratios for Shingrix® vaccine uptake for each potential uptake-related factor and 95%CI.

Model 1: Minimally adjusted, where each covariate is included individually adjusted with age and sex; Model 2: Model 1 + prior zoster vaccination; Model 3: Model 2 + region + IMD + ethnicity; Model 4: Model 3 + immunosuppression risk level; Model 5: Model 4 + clinical factors (CKD, COPD, DM, Dementia, CMD, SMI); Model 6: Model 5 + lifestyle factors (harmful alcohol use, smoking, overweight/obese); Model 7: Model 6 + living alone + care home residence.

| | | Model 1 Minimally adjusted (adjusted for age and sex) N=86,197 | Model 2 (adjusted for variables in Model 1 + previous zoster vaccination) N=86,197 | Model 3 (adjusted for variables in Model 2 + region+IMD+ ethnicity) N=69,260 | Model 4 (adjusted for variables in Model 3 + immuno-suppression risk level) N=69,260 | Model 5 (adjusted for variables in Model 4 + clinical factors) N=69,260 | Model 6 (Main model) (adjusted for variables in Model 5 + lifestyle factors) N=60,305 | Model 7 (Full model) (adjusted for variables in Model 6 + living alone + care home residence) N=60,305 |
|---|---|---|---|---|---|---|---|---|
| **Age (years)** | 70 | ref | ref | ref | ref | ref | ref | ref |
| | 71 | 1.06 (1.00, 1.13) | 1.06 (1.00, 1.13) | 1.08 (1.01, 1.16) | 1.09 (1.01, 1.16) | 1.09 (1.02, 1.17) | 1.10 (1.02, 1.18) | 1.10 (1.02, 1.18) |
| | 72 | 0.85 (0.79, 0.91) | 0.85 (0.79, 0.91) | 0.89 (0.82, 0.95) | 0.89 (0.83, 0.96) | 0.89 (0.83, 0.96) | 0.89 (0.82, 0.96) | 0.89 (0.82, 0.96) |
| | 73 | 0.70 (0.65, 0.75) | 0.70 (0.65, 0.75) | 0.69 (0.63, 0.75) | 0.69 (0.63, 0.75) | 0.69 (0.63, 0.76) | 0.70 (0.64, 0.77) | 0.70 (0.64, 0.77) |
| | 74 | 0.65 (0.60, 0.70) | 0.65 (0.60, 0.70) | 0.63 (0.58, 0.69) | 0.63 (0.58, 0.69) | 0.63 (0.58, 0.69) | 0.65 (0.59, 0.71) | 0.65 (0.59, 0.71) |
| | 75 | 0.61 (0.56, 0.66) | 0.61 (0.56, 0.66) | 0.62 (0.56, 0.68) | 0.62 (0.56, 0.69) | 0.62 (0.57, 0.69) | 0.62 (0.55, 0.68) | 0.62 (0.56, 0.69) |
| | 76 | 0.50 (0.45, 0.56) | 0.50 (0.45, 0.56) | 0.48 (0.43, 0.54) | 0.48 (0.43, 0.54) | 0.49 (0.43, 0.55) | 0.49 (0.43, 0.56) | 0.49 (0.44, 0.56) |
| | 77 | 0.43 (0.38, 0.48) | 0.43 (0.38, 0.48) | 0.42 (0.37, 0.48) | 0.42 (0.37, 0.48) | 0.42 (0.37, 0.48) | 0.42 (0.36, 0.48) | 0.42 (0.36, 0.48) |
| | 78 | 0.41 (0.37, 0.47) | 0.41 (0.37, 0.47) | 0.40 (0.35, 0.47) | 0.40 (0.35, 0.47) | 0.41 (0.35, 0.47) | 0.41 (0.35, 0.48) | 0.41 (0.35, 0.48) |
| | 79 | 0.72 (0.66, 0.79) | 0.72 (0.66, 0.79) | 0.73 (0.66, 0.81) | 0.74 (0.67, 0.81) | 0.75 (0.68, 0.83) | 0.75 (0.67, 0.83) | 0.75 (0.68, 0.84) |
| **Sex** | Male | ref | ref | ref | ref | ref | ref | ref |
| | Female | 0.96 (0.93, 1.00) | 0.96 (0.93, 1.00) | 0.97 (0.93, 1.01) | 0.94 (0.90, 0.98) | 0.94 (0.90, 0.98) | 0.94 (0.90, 0.98) | 0.93 (0.89, 0.98) |
| **Zoster infection** | No | ref | ref | ref | ref | ref | ref | ref |

| | | | | | | | | |
|---|---|---|---|---|---|---|---|---|
| **in the last year** | Yes | 1.35 (1.15, 1.58) | 1.35 (1.15, 1.58) | 1.37 (1.15, 1.64) | 1.36 (1.14, 1.62) | 1.36 (1.14, 1.63) | 1.38 (1.14, 1.65) | 1.37 (1.14, 1.65) |
| **Region** | London | ref | ref | ref | ref | ref | ref | ref |
| | East Midlands | 1.17 (1.01, 1.35) | 1.17 (1.01, 1.35) | 1.09 (0.80, 1.48) | 1.07 (0.79, 1.45) | 1.07 (0.79, 1.45) | 1.08 (0.82, 1.42) | 1.07 (0.82, 1.41) |
| | East of England | 1.52 (1.38, 1.69) | 1.52 (1.37, 1.68) | 1.27 (1.03, 1.55) | 1.25 (1.02, 1.54) | 1.23 (1.00, 1.52) | 1.22 (0.99, 1.50) | 1.21 (0.98, 1.50) |
| | North East | 1.12 (1.01, 1.24) | 1.12 (1.01, 1.24) | 1.04 (0.86, 1.26) | 1.02 (0.84, 1.24) | 1.01 (0.83, 1.23) | 1.00 (0.82, 1.23) | 1.00 (0.82, 1.23) |
| | North West | 0.97 (0.91,1.04) | 0.97 (0.90, 1.04) | 0.91 (0.79, 1.04) | 0.90 (0.79, 1.03) | 0.90 (0.79, 1.02) | 0.88 (0.77, 1.01) | 0.88 (0.77, 1.01) |
| | South East | 1.29 (1.21, 1.38) | 1.29 (1.21, 1.37) | 1.10 (0.97, 1.24) | 1.09 (0.96, 1.24) | 1.08 (0.95, 1.23) | 1.07 (0.94, 1.21) | 1.07 (0.94, 1.21) |
| | South West | 1.27 (1.18, 1.37) | 1.27 (1.18, 1.37) | 1.15 (0.99, 1.33) | 1.15 (0.98, 1.34) | 1.12 (0.97, 1.31) | 1.11 (0.95, 1.29) | 1.11 (0.95, 1.29) |
| | West Midlands | 1.23 (1.15, 1.32) | 1.23 (1.15, 1.32) | 1.12 (0.98, 1.28) | 1.11 (0.97, 1.27) | 1.09 (0.95, 1.26) | 1.09 (0.95, 1.25) | 1.09 (0.95, 1.25) |
| | Yorkshire and The Humber | 1.09 (0.97, 1.21) | 1.08 (0.97, 1.21) | 0.95 (0.74, 1.22) | 0.95 (0.74, 1.22) | 0.94 (0.73, 1.21) | 0.95 (0.73, 1.23) | 0.95 (0.73, 1.23) |
| **Index of multiple deprivation quintile** | 1 (Least deprived) | ref | ref | ref | ref | ref | ref | ref |
| | 2 | 0.86 (0.80, 0.92) | 0.86 (0.81, 0.91) | 0.87 (0.81, 0.93) | 0.86 (0.81, 0.92) | 0.87 (0.82, 0.93) | 0.87 (0.81, 0.93) | 0.87 (0.81, 0.93) |
| | 3 | 0.78 (0.73, 0.84) | 0.79 (0.75, 0.84) | 0.79 (0.74, 0.85) | 0.79 (0.73, 0.85) | 0.80 (0.74, 0.86) | 0.80 (0.74, 0.86) | 0.80 (0.74, 0.86) |
| | 4 | 0.65 (0.60, 0.71) | 0.65 (0.60, 0.71) | 0.68 (0.62, 0.74) | 0.67 (0.62, 0.73) | 0.69 (0.64, 0.75) | 0.69 (0.63, 0.75) | 0.69 (0.63, 0.75) |
| | 5 (Most deprived) | 0.55 (0.50, 0.60) | 0.55 (0.50, 0.60) | 0.59 (0.54, 0.65) | 0.58 (0.53, 0.63) | 0.61 (0.56, 0.67) | 0.61 (0.56, 0.67) | 0.61 (0.56, 0.67) |
| **Ethnicity** | 0. White | ref | ref | ref | ref | ref | ref | ref |
| | 1. South Asian | 0.70 (0.62, 0.78) | 0.70 (0.62, 0.78) | 0.78 (0.68, 0.91) | 0.76 (0.66, 0.89) | 0.74 (0.64, 0.86) | 0.72 (0.62, 0.83) | 0.72 (0.62, 0.83) |
| | 2. Black | 0.48 (0.40, 0.58) | 0.48 (0.40, 0.58) | 0.57 (0.46, 0.71) | 0.56 (0.46, 0.70) | 0.54 (0.43, 0.67) | 0.53 (0.43, 0.66) | 0.53 (0.43, 0.66) |
| | 3. Other/Mixed | 0.79 (0.71, 0.88) | 0.79 (0.71, 0.88) | 0.81 (0.69, 0.95) | 0.80 (0.69, 0.93) | 0.78 (0.67, 0.91) | 0.75 (0.65, 0.86) | 0.75 (0.65, 0.86) |
| **Immunosuppres** | High | ref | ref | ref | ref | ref | ref | ref |

| | | | | | | | | |
|---|---|---|---|---|---|---|---|---|
| sion risk level | Low | 1.47 (1.42, 1.52) | 1.47 (1.41, 1.52) | 1.45 (1.38, 1.53) | 1.45 (1.38, 1.53) | 1.49 (1.41, 1.57) | 1.48 (1.40, 1.56) | 1.48 (1.40, 1.57) |
| **Common mental disorders (CMD)** | No | ref | ref | ref | ref | ref | ref | ref |
| | Yes | 0.88 (0.84, 0.92) | 0.88 (0.84, 0.92) | 0.91 (0.87, 0.96) | 0.90 (0.86, 0.95) | 0.93 (0.89, 0.98) | 0.93 (0.88, 0.98) | 0.93 (0.89, 0.98) |
| **Severe mental illness (SMI)** | No | ref | ref | ref | ref | ref | ref | ref |
| | Yes | 0.56 (0.46, 0.68) | 0.56 (0.46, 0.68) | 0.61 (0.48, 0.76) | 0.62 (0.49, 0.78) | 0.66 (0.53, 0.83) | 0.67 (0.53, 0.84) | 0.72 (0.57, 0.91) |
| **Dementia** | No | ref | ref | ref | ref | ref | ref | ref |
| | Yes | 0.60 (0.51, 0.70) | 0.60 (0.51, 0.70) | 0.64 (0.54, 0.76) | 0.63 (0.53, 0.75) | 0.66 (0.55, 0.78) | 0.71 (0.60, 0.84) | 0.79 (0.66, 0.94) |
| **Diabetes Mellitus** | No | ref | ref | ref | ref | ref | ref | |
| | Yes | 0.93 (0.89, 0.96) | 0.93 (0.89, 0.96) | 1.00 (0.95, 1.05) | 0.99 (0.95, 1.04) | 1.02 (0.97, 1.07) | 0.99 (0.94, 1.04) | 0.99 (0.94, 1.04) |
| **Chronic obstructive pulmonary disease (COPD)** | No | ref | ref | ref | ref | ref | ref | |
| | Yes | 0.79 (0.75, 0.83) | 0.79 (0.75, 0.83) | 0.83 (0.78, 0.87) | 0.77 (0.73, 0.82) | 0.78 (0.74, 0.83) | 0.79 (0.75, 0.84) | 0.79 (0.74, 0.84) |
| **Chronic kidney disease (CKD)** | No | ref | ref | ref | ref | ref | ref | |
| | Yes | 0.92 (0.88, 0.97) | 0.92 (0.88, 0.97) | 0.94 (0.89, 1.00) | 0.97 (0.92, 1.03) | 0.98 (0.92, 1.03) | 0.95 (0.90, 1.01) | 0.95 (0.90, 1.01) |
| **Harmful alcohol use** | No | ref | ref | ref | ref | ref | ref | |
| | Yes | 0.65 (0.56, 0.75) | 0.65 (0.56, 0.75) | 0.68 (0.57, 0.80) | 0.69 (0.58, 0.81) | 0.74 (0.62, 0.87) | 0.74 (0.62, 0.89) | 0.76 (0.64, 0.90) |
| **Smoking history** | No | ref | ref | ref | ref | ref | ref | |
| | Yes | 0.88 (0.85, 0.92) | 0.88 (0.85, 0.92) | 0.93 (0.88, 0.98) | 0.92 (0.87, 0.97) | 0.96 (0.91, 1.01) | 0.97 (0.91, 1.02) | 0.96 (0.91, 1.02) |
| **Overweight/obese** | BMI <25 kg/m$^2$ | ref | ref | ref | ref | ref | ref | |
| | BMI ≥25 kg/m$^2$ | 1.12 (1.08, 1.17) | 1.12 (1.08, 1.17) | 1.13 (1.08, 1.18) | 1.12 (1.07, 1.17) | 1.11 (1.06, 1.16) | 1.11 (1.06, 1.17) | 1.11 (1.06, 1.16) |
| **Living alone** | No | ref | ref | ref | ref | ref | ref | |
| | Yes | 0.79 (0.72, 0.87) | 0.79 (0.72, 0.87) | 0.91 (0.82, 1.01) | 0.89 (0.80, 0.99) | 0.98 (0.88, 1.09) | 1.00 (0.89, 1.11) | 0.99 (0.89, 1.11) |
| | No | ref | ref | ref | ref | ref | ref | |

| | | | | | | | | |
|---|---|---|---|---|---|---|---|---|
| **Care home residence** | Yes | 0.32 (0.25, 0.41) | 0.32 (0.25, 0.41) | 0.37 (0.26, 0.51) | 0.36 (0.26, 0.50) | 0.42 (0.30, 0.59) | 0.44 (0.31, 0.62) | 0.44 (0.31, 0.62) |

Supplementary Table S7: Sensitivity analysis, change in number from risk at entry to the study to highest risk level/category over time under study.

| | | Immunosuppression risk level at entry | | | Immunosuppression risk level at highest point during study | | |
|---|---|---|---|---|---|---|---|
| | | N (% of total study population) | Vaccine uptake (%) | Median follow-up time [IQR] | N (% of total study population) | Vaccine uptake (%) | Median follow-up time [IQR] |
| **Immunosuppression risk level (2 categories)** | **High** | 37,145 (43.1%) | 14.3% | 299 [95-483] | 38,931 (45.2%) | 14.6% | 303 [101-520] |
| | **Low** | 49,052 (56.9%) | 19.6% | 426 [110-729] | 47,266 (54.8%) | 19.6% | 426 [106-729] |
| **Immunosuppression risk level (5 categories)** | **1 (Highest risk)** | 9,442 (11.0%) | 18.4% | 426 [176-729] | 10,038 (11.6%) | 18.5% | 426 [189-729] |
| | **2** | 2,009 (2.3%) | 19.1% | 668 [274-729] | 2,009 (2.3%) | 19.1% | 688 [272-729] |
| | **3** | 25,694 (29.8%) | 12.4% | 243 [77-391] | 26,884 (31.2%) | 12.8% | 258 [82-419] |
| | **4** | 28,359 (32.9%) | 17.8% | 301 [92-669] | 29,292 (34.0%) | 18.4% | 303 [92-729] |
| | **5 (Lowest risk)** | 20,693 (24.0%) | 22.1% | 668 [303-729] | 17,974 (20.9%) | 21.5% | 668 [303-729] |

The two immunosuppression risk level categories were defined as: higher (bone marrow compromised, solid organ transplant) and lower risk (recently treated cancer, non-cancer immunosuppressives, immunodeficiency). The five immunosuppression risk level categories were defined as: 1) bone marrow compromised; 2) solid organ transplant; 3) recently treated cancer; 4) non-cancer related immunosuppressives; and 5) immunodeficiency (primary and acquired).

Supplementary Table S8: Results of sensitivity analyses: odds ratios for Shingrix® uptake (95% confidence intervals)

| | | **Main analysis results** (adjusted for age, sex, Zoster infection in the last year, region, ethnicity, IMD, immunosuppression risk category, clinical and lifestyle factors) (N=60,305) | **Sensitivity analysis 1:** Static cohort (N=30,782) | **Sensitivity analysis 2:** Time updated covariates (N=60,305) | **Sensitivity analysis 3:** Restricted to at least 13 months follow-up (n=43,986) | **Sensitivity analysis 4:** Age 70 only (n=27,688) | **Sensitivity analysis 5:** More granular ethnicity categories (N=60,305) | **Sensitivity analysis 6:** More granular immunosuppression risk-level categories (N=60,305) |
|---|---|---|---|---|---|---|---|---|
| **Age (years)** | 70 | ref | ref | ref | ref | - | ref | ref |
| | 71 | 1.10 (1.02, 1.18) | 0.79 (0.72, 0.87) | 1.10 (1.03, 1.19) | 0.78 (0.72, 0.84) | - | 1.10 (1.02, 1.18) | 1.11 (1.03, 1.20) |
| | 72 | 0.89 (0.82, 0.96) | 0.62 (0.57, 0.69) | 0.89 (0.83, 0.97) | 0.63 (0.58, 0.69) | - | 0.88 (0.82, 0.95) | 0.89 (0.82, 0.96) |
| | 73 | 0.70 (0.64, 0.77) | 0.49 (0.44, 0.55) | 0.70 (0.64, 0.77) | 0.50 (0.46, 0.56) | - | 0.70 (0.64, 0.77) | 0.70 (0.64, 0.77) |
| | 74 | 0.65 (0.59, 0.71) | 0.46 (0.41, 0.52) | 0.65 (0.60, 0.71) | 0.46 (0.42, 0.51) | - | 0.65 (0.59, 0.71) | 0.64 (0.59, 0.71) |
| | 75 | 0.62 (0.55, 0.68) | 0.46 (0.41, 0.52) | 0.62 (0.56, 0.69) | 0.44 (0.40, 0.49) | - | 0.62 (0.55, 0.68) | 0.61 (0.55, 0.68) |
| | 76 | 0.49 (0.43, 0.56) | 0.35 (0.30, 0.41) | 0.49 (0.44, 0.56) | 0.35 (0.31, 0.40) | - | 0.49 (0.43, 0.56) | 0.49 (0.43, 0.55) |
| | 77 | 0.42 (0.36, 0.48) | 0.31 (0.27, 0.37) | 0.43 (0.37, 0.49) | 0.30 (0.26, 0.35) | - | 0.42 (0.36, 0.48) | 0.41 (0.36, 0.48) |
| | 78 | 0.41 (0.35, 0.48) | 0.28 (0.24, 0.34) | 0.41 (0.36, 0.48) | 0.29 (0.25, 0.35) | - | 0.41 (0.35, 0.48) | 0.41 (0.35, 0.48) |
| | 79 | 0.75 (0.67, 0.83) | 0.52 (0.46, 0.59) | 0.73 (0.65, 0.81) | 0.07 (0.04, 0.14) | - | 0.75 (0.67, 0.83) | 0.75 (0.68, 0.84) |
| **Sex** | Male | ref | ref | ref | ref | ref | ref | ref |

| | | | | | | | | |
|---|---|---|---|---|---|---|---|---|
| | Female | 0.94 (0.90, 0.98) | 0.88 (0.83, 0.93) | 0.94 (0.90, 0.98) | 0.93 (0.88, 0.97) | 1.04 (0.97, 1.12) | 0.93 (0.89, 0.98) | 0.95 (0.91, 0.99) |
| **Zoster infection in the last year** | No | ref | ref | ref | ref | ref | ref | ref |
| | Yes | 1.38 (1.14, 1.65) | 1.40 (1.11, 1.77) | 1.33 (1.10, 1.60) | 1.49 (1.22, 1.82) | 1.16 (0.83, 1.61) | 1.37 (1.14, 1.65) | 1.35 (1.12, 1.63) |
| **Region** | London | ref | ref | ref | ref | ref | ref | ref |
| | East Midlands | 1.08 (0.82, 1.42) | 1.04 (0.68, 1.58) | 1.07 (0.81, 1.42) | 1.27 (1.01, 1.58) | 0.88 (0.64, 1.21) | 1.09 (0.82, 1.43) | 1.11 (0.84, 1.46) |
| | East of England | 1.22 (0.99, 1.50) | 1.21 (0.96, 1.52) | 1.23 (0.99, 1.52) | 1.17 (0.93, 1.46) | 1.38 (1.04, 1.82) | 1.23 (1.00, 1.51) | 1.24 (1.01, 1.53) |
| | North East | 1.00 (0.82, 1.23) | 0.95 (0.75, 1.22) | 1.00 (0.82, 1.23) | 1.01 (0.82, 1.25) | 1.19 (0.87, 1.62) | 1.01 (0.82, 1.24) | 1.02 (0.83, 1.25) |
| | North West | 0.88 (0.77, 1.01) | 0.88 (0.75, 1.04) | 0.89 (0.78, 1.01) | 0.84 (0.73, 0.97) | 0.91 (0.76, 1.09) | 0.89 (0.78, 1.02) | 0.90 (0.78, 1.02) |
| | South East | 1.07 (0.94, 1.21) | 1.02 (0.87, 1.19) | 1.07 (0.95, 1.22) | 1.05 (0.92, 1.21) | 1.09 (0.92, 1.30) | 1.08 (0.95, 1.22) | 1.09 (0.96, 1.24) |
| | South West | 1.11 (0.95, 1.29) | 1.03 (0.86, 1.23) | 1.11 (0.95, 1.30) | 1.12 (0.95, 1.32) | 1.19 (0.99, 1.45) | 1.12 (0.96, 1.31) | 1.13 (0.97, 1.32) |
| | West Midlands | 1.09 (0.95, 1.25) | 1.08 (0.92, 1.28) | 1.10 (0.95, 1.26) | 1.06 (0.91, 1.23) | 1.14 (0.96, 1.36) | 1.10 (0.95, 1.26) | 1.11 (0.97, 1.27) |
| | Yorkshire and The Humber | 0.95 (0.73, 1.23) | 0.84 (0.63, 1.14) | 0.95 (0.73, 1.23) | 0.96 (0.70, 1.32) | 1.06 (0.74, 1.50) | 0.95 (0.73, 1.24) | 0.96 (0.74, 1.24) |
| **Index of multiple deprivation quintiles** | 1 (Least deprived) | ref | ref | ref | ref | ref | | ref |
| | 2 | 0.87 (0.81, 0.93) | 0.85 (0.78, 0.93) | 0.86 (0.80, 0.92) | 0.84 (0.78, 0.91) | 0.88 (0.79, 0.98) | 0.87 (0.81, 0.93) | 0.87 (0.81, 0.93) |
| | 3 | 0.80 (0.74, 0.86) | 0.79 (0.72, 0.87) | 0.79 (0.73, 0.85) | 0.79 (0.72, 0.85) | 0.81 (0.72, 0.91) | 0.80 (0.74, 0.86) | 0.80 (0.74, 0.86) |
| | 4 | 0.69 (0.63, 0.75) | 0.70 (0.63, 0.78) | 0.68 (0.62, 0.74) | 0.69 (0.63, 0.75) | 0.66 (0.58, 0.75) | 0.69 (0.63, 0.76) | 0.69 (0.63, 0.75) |
| | 5 (Most deprived) | 0.61 (0.56, 0.67) | 0.63 (0.56, 0.70) | 0.59 (0.54, 0.65) | 0.60 (0.54, 0.67) | 0.57 (0.50, 0.65) | 0.61 (0.56, 0.67) | 0.61 (0.55, 0.67) |
| **Ethnicity (5** | 0. White | ref | ref | ref | ref | ref | - | ref |

| | | | | | | | | |
|---|---|---|---|---|---|---|---|---|
| **categories)** | 1. South Asian | 0.72 (0.62, 0.83) | 0.76 (0.63, 0.91) | 0.72 (0.61, 0.84) | 0.72 (0.62, 0.85) | 0.80 (0.65, 0.97) | - | 0.70 (0.60, 0.81) |
| | 2. Black | 0.53 (0.43, 0.66) | 0.54 (0.40, 0.73) | 0.53 (0.42, 0.65) | 0.53 (0.42, 0.67) | 0.50 (0.36, 0.69) | - | 0.50 (0.40, 0.62) |
| | 3. Other/Mixed | 0.75 (0.65, 0.86) | 0.87 (0.72, 1.04) | 0.77 (0.65, 0.90) | 0.76 (0.65, 0.88) | 0.67 (0.54, 0.83) | - | 0.74 (0.64, 0.85) |
| **Ethnicity (16 categories)** | 01. British | - | - | - | - | - | ref | - |
| | 02. Irish | - | - | - | - | - | 0.99 (0.77, 1.27) | - |
| | 03. Other White | - | - | - | - | - | 1.03 (0.94, 1.14) | - |
| | 04. Indian | - | - | - | - | - | 0.88 (0.71, 1.09) | - |
| | 05. Pakistani | - | - | - | - | - | 0.46 (0.31, 0.68) | - |
| | 06. Bangladeshi | - | - | - | - | - | 0.68 (0.36, 1.28) | - |
| | 07. Other Asian | - | - | - | - | - | 0.68 (0.52, 0.87) | - |
| | 08. Caribbean | - | - | - | - | - | 0.57 (0.41, 0.78) | - |
| | 09. African | - | - | - | - | - | 0.56 (0.40, 0.78) | - |
| | 10. Other Black | - | - | - | - | - | 0.43 (0.26, 0.71) | - |
| | 11. White and Black Caribbean | - | - | - | - | - | 0.33 (0.08, 1.36) | - |
| | 12. White and Black African | - | - | - | - | - | 0.43 (0.10, 1.96) | - |
| | 13. White and Asian | - | - | - | - | - | 1.05 (0.43, 2.52) | - |
| | 14. Chinese | - | - | - | - | - | 0.69 (0.36, 1.30) | - |
| | 15. Other ethnic group | - | - | - | - | - | 0.71 (0.48, 1.07) | - |
| | 16. Other Mixed | - | - | - | - | - | 0.77 (0.66, 0.90) | - |
| **Immunosuppression risk level (2 category)** | High | ref | ref | ref | ref | ref | ref | - |
| | Low | 1.48 (1.40, 1.56) | 1.47 (1.37, 1.58) | 1.42 (1.35, 1.50) | 1.40 (1.32, 1.49) | 1.33 (1.23, 1.44) | 1.48 (1.40, 1.56) | - |
| **Immunosuppression risk level (5 category)** | 1 (Highest risk) | – | – | – | – | - | - | ref |
| | 2 | – | – | – | – | - | - | 1.05 (0.90, 1.23) |
| | 3 | – | – | – | – | - | - | 0.61 (0.56, 0.67) |

| | | | | | | | | |
|---|---|---|---|---|---|---|---|---|
| | 4 | – | – | – | – | - | - | 0.93 (0.86, 1.01) |
| | 5 (Lowest risk) | – | – | – | – | - | - | 1.27 (1.17, 1.37) |
| **Common mental disorders** | No | ref | ref | ref | ref | ref | ref | ref |
| | Yes | 0.93 (0.88, 0.98) | 0.97 (0.91, 1.04) | 0.95 (0.91, 1.00) | 0.95 (0.90, 1.01) | 0.87 (0.80, 0.94) | 0.93 (0.88, 0.97) | 0.93 (0.88, 0.98) |
| **Severe mental illness** | No | ref | | | | | ref | ref |
| | Yes | 0.67 (0.53, 0.84) | 0.67 (0.50, 0.89) | 0.62 (0.50, 0.78) | 0.71 (0.55, 0.92) | 0.84 (0.59, 1.18) | 0.67 (0.53, 0.84) | 0.66 (0.53, 0.83) |
| **Dementia** | No | ref | ref | ref | ref | ref | ref | ref |
| | Yes | 0.71 (0.60, 0.84) | 0.77 (0.62, 0.95) | 0.85 (0.74, 0.98) | 0.73 (0.59, 0.90) | 0.76 (0.52, 1.10) | 0.71 (0.60, 0.85) | 0.69 (0.58, 0.82) |
| **Diabetes Mellitus** | No | ref | ref | ref | ref | ref | ref | ref |
| | Yes | 0.99 (0.94, 1.04) | 1.01 (0.95, 1.07) | 1.06 (1.01, 1.12) | 1.02 (0.97, 1.08) | 0.99 (0.92, 1.07) | 0.99 (0.94, 1.04) | 0.99 (0.95, 1.05) |
| **Chronic obstructive pulmonary disease** | No | ref | ref | ref | ref | ref | ref | ref |
| | Yes | 0.79 (0.75, 0.84) | 0.83 (0.77, 0.90) | 0.84 (0.79, 0.89) | 0.79 (0.74, 0.84) | 0.93 (0.85, 1.02) | 0.79 (0.75, 0.84) | 0.84 (0.79, 0.89) |
| **Chronic kidney disease** | No | ref | ref | ref | ref | ref | ref | ref |
| | Yes | 0.95 (0.90, 1.01) | 0.92 (0.85, 1.00) | 1.01 (0.95, 1.07) | 0.96 (0.90, 1.03) | 0.91 (0.82, 1.01) | 0.96 (0.90, 1.02) | 0.91 (0.85, 0.96) |
| **Harmful alcohol use** | No | ref | ref | ref | ref | ref | ref | ref |
| | Yes | 0.74 (0.62, 0.89) | 0.76 (0.59, 0.97) | 0.76 (0.64, 0.90) | 0.75 (0.61, 0.91) | 0.72 (0.55, 0.95) | 0.76 (0.64, 0.90) | 0.74 (0.62, 0.89) |
| **Smoking history** | No | ref | ref | ref | ref | ref | ref | ref |
| | Yes | 0.97 (0.91, 1.02) | 0.94 (0.88, 1.01) | 0.97 (0.92, 1.03) | 0.97 (0.91, 1.03) | 0.92 (0.84, 1.01) | 0.97 (0.92, 1.03) | 0.98 (0.93, 1.03) |
| **Overweight/obese** | BMI <25 kg/m$^2$ | ref | ref | ref | ref | ref | ref | ref |
| | BMI ≥25 kg/m$^2$ | 1.11 (1.06, 1.17) | 1.08 (1.02, 1.15) | 1.09 (1.04, 1.15) | 1.06 (1.01, 1.11) | 1.17 (1.08, 1.26) | 1.12 (1.07, 1.17) | 1.12 (1.07, 1.18) |
| **Living alone** | No | ref | ref | ref | ref | ref | ref | ref |
| | Yes | 1.00 (0.89, 1.11) | 0.99 (0.86, 1.12) | 1.03 (0.93, 1.14) | 1.02 (0.90, 1.15) | 1.01 (0.84, 1.21) | 0.99 (0.89, 1.11)) | 0.99 (0.89, 1.10) |
| | No | ref | ref | ref | ref | ref | ref | ref |

| | | | | | | | | |
|---|---|---|---|---|---|---|---|---|
| **Care home residence** | Yes | 0.44 (0.31, 0.62) | 0.42 (0.26, 0.68) | 0.48 (0.37, 0.64) | 0.54 (0.35, 0.82) | 0.59 (0.34, 1.04) | 0.44 (0.31, 0.62) | 0.44 (0.31, 0.62) |

Supplementary Table S9: Percentage of Shingrix® uptake by ethnicity. Number and percentages of individuals in the dataset by ethnicity, with corresponding vaccination numbers and percentages within each group.

| Ethnicity (4 groups) | N (% of total study population) | Vaccine uptake (%) | Median follow-up time [IQR] | Ethnicity (16 groups) | N (% of total study population)% column | Vaccine uptake (%) | Median follow-up time [IQR] |
|---|---|---|---|---|---|---|---|
| **White** | 76,007 (88.2%) | 18.0% | 352 [106-729] | British | 70,492 (81.8%) | 18.0% | 355 [107-729] |
| | | | | Irish | 696 (0.8%) | 16.4% | 330 [95-714] |
| | | | | Other White | 4,819 (5.6%) | 17.6% | 332 [102-704] |
| **South Asian** | 2,579 (3.0%) | 14.2% | 365 [92-729] | Indian | 1,103 (1.3%) | 17.5% | 365 [111-729] |
| | | | | Pakistani | 573 (0.7%) | 8.0% | 402 [93-729] |
| | | | | Bangladeshi | 152 (0.2%) | 12.5% | 426 [146-684] |
| | | | | Other Asian | 751 (0.9%) | 14.5% | 365 [103-729] |
| **Black** | 1,205 (1.4%) | 10.0% | 374 [94-729] | Caribbean | 452 (0.5%) | 9.7% | 413 [96-729] |
| | | | | African | 498 (0.6%) | 11.0% | 303 [62-630] |
| | | | | Other Black | 255 (0.3%) | 8.6% | 365 [90-729] |
| **Other/Mixed** | 2,705 (3.1%) | 15.1% | 365 [99-729] | White and Black Caribbean | 37 (<0.0%) | <13.5%* | 412 [101-729] |
| | | | | White and Black African | 34 (<0.1%) | <14.7%* | 365 [94-729] |
| | | | | White and Asian | 39 (<0.1%) | 23.1% | 426 [93-729] |
| | | | | Chinese | 150 (0.2%) | 12.7% | 370 [98-729] |
| | | | | Other ethnic group | 360 (0.4%) | 15.3% | 304 [61-681] |
| | | | | Other Mixed | 2,085 (2.4%) | 15.3% | 350 [96-602] |
| ***Missing*** | 3,701 (4.3%) | 10.1% | 287 [92-577] | Missing | 3,701 (4.3%) | 10.1% | 287 [92-577] |

*Counts less than 5 are redacted.

#### Supplementary Table S10: Characteristics of individuals with at least 13 months of available follow up by one- and two-dose Shingrix(R) uptake

Individuals with at least 13 months follow up, N=50,486 (58.6% of the original cohort).

| | | No vaccination (N=38,386) | | One dose vaccine only (N=6,590) | | Both first and second doses (N=5,510) | |
|---|---|---|---|---|---|---|---|
| | | N (% of those not vaccinated) | Median follow-up time (days) [IQR] | N (% of those with only one dose) | Median follow-up time (days) [IQR] | N (% of those with two doses) | Median follow-up time (days) [IQR] |
| **Age (years)** | 70 | 10,439 (27.2%) | 426 [347 - 675] | 2,726 (41.4%) | 426 [347 - 675] | 2,002 (36.3%) | 426 [426 - 729] |
| | 71 | 5,436 (14.2%) | 641 [365 - 729] | 1,186 (18.0%) | 641 [365 - 729] | 897 (16.3%) | 729 [729 - 729] |
| | 72 | 4,549 (11.9%) | 704 [365 - 729] | 737 (11.2%) | 704 [365 - 729] | 674 (12.2%) | 729 [729 - 729] |
| | 73 | 4,166 (10.9%) | 724 [404 - 729] | 544 (8.3%) | 724 [404 - 729] | 525 (9.5%) | 729 [729 - 729] |
| | 74 | 4,098 (10.7%) | 729 [407 - 729] | 494 (7.5%) | 729 [407 - 729] | 472 (8.6%) | 729 [729 - 729] |
| | 75 | 3,334 (8.7%) | 729 [409 - 729] | 381 (5.8%) | 729 [409 - 729] | 376 (6.8%) | 729 [729 - 729] |
| | 76 | 2,327 (6.1%) | 729 [429 - 729] | 212 (3.2%) | 729 [429 - 729] | 240 (4.4%) | 729 [729 - 729] |
| | 77 | 2,225 (5.8%) | 729 [415 - 729] | 166 (2.5%) | 729 [415 - 729] | 192 (3.5%) | 729 [729 - 729] |
| | 78 | 1,812 (4.7%) | 668 [410 - 668] | 144 (2.2%) | 668 [410 - 668] | 132 (2.4%) | 668 [668 - 668] |
| **Sex** | Male | 15,642 (40.7%) | 566 [365 - 729] | 2,875 (43.6%) | 566 [365 - 729] | 2,343 (42.5%) | 729 [516 - 729] |
| | Female | 22,744 (59.3%) | 625 [372 - 729] | 3,715 (56.4%) | 625 [372 - 729] | 3,167 (57.5%) | 729 [524 - 729] |
| **Zoster infection in the last year** | No | 37,995 (99.0%) | 597 [366 - 729] | 6,515 (98.9%) | 597 [366 - 729] | 5,423 (98.4%) | 729 [520 - 729] |
| | Yes | 391 (1.0%) | 650 [365 - 729] | 75 (1.1%) | 650 [365 - 729] | 87 (1.6%) | 729 [496 - 729] |
| **Region** | London | 4,518 (11.8%) | 604 [396 - 729] | 795 (12.1%) | 604 [396 - 729] | 483 (8.8%) | 729 [446 - 729] |
| | East Midlands | 550 (1.4%) | 518 [365 - 729] | 105 (1.6%) | 518 [365 - 729] | 93 (1.7%) | 729 [426 - 729] |
| | East of England | 1,381 (3.6%) | 593 [365 - 729] | 279 (4.2%) | 593 [365 - 729] | 273 (5.0%) | 729 [540 - 729] |
| | North East | 1,546 (4.0%) | 587 [365 - 729] | 263 (4.0%) | 587 [365 - 729] | 240 (4.4%) | 729 [467 - 729] |
| | North West | 9,159 (23.9%) | 594 [371 - 729] | 1,347 (20.4%) | 594 [371 - 729] | 1,012 (18.4%) | 729 [562 - 729] |
| | South East | 8,621 (22.5%) | 602 [365 - 729] | 1,497 (22.7%) | 602 [365 - 729] | 1,492 (27.1%) | 729 [508 - 729] |
| | South West | 4,497 (11.7%) | 603 [374 - 729] | 827 (12.5%) | 603 [374 - 729] | 738 (13.4%) | 729 [525 - 729] |
| | West Midlands | 6,994 (18.2%) | 604 [365 - 729] | 1,295 (19.7%) | 604 [365 - 729] | 1,023 (18.6%) | 729 [569 - 729] |
| | Yorkshire and The Humber | 1,120 (2.9%) | 584 [377 - 729] | 182 (2.8%) | 584 [377 - 729] | 156 (2.8%) | 729 [440 - 729] |
| **Index of multiple deprivation quintile** | 1 (Least deprived) | 6,801 (17.7%) | 620 [367 - 729] | 1,295 (19.7%) | 620 [367 - 729] | 1,383 (25.1%) | 729 [535 - 729] |
| | 2 | 7,061 (18.4%) | 604 [365 - 729] | 1,239 (18.8%) | 604 [365 - 729] | 1,129 (20.5%) | 729 [556 - 729] |
| | 3 | 6,303 (16.4%) | 602 [377 - 729] | 1,088 (16.5%) | 602 [377 - 729] | 914 (16.6%) | 729 [492 - 729] |
| | 4 | 5,953 (15.5%) | 595 [369 - 729] | 954 (14.5%) | 595 [369 - 729] | 695 (12.6%) | 729 [494 - 729] |

| | | | | | | | |
|---|---|---|---|---|---|---|---|
| | 5 (Most deprived) | 5,313 (13.8%) | 576 [382 - 729] | 772 (11.7%) | 576 [382 - 729] | 473 (8.6%) | 729 [518 - 729] |
| | NA | 6,955 (18.1%) | 591 [365 - 729] | 1,242 (18.8%) | 591 [365 - 729] | 916 (16.6%) | 729 [490 - 729] |
| **Ethnicity** | 0. White | 33,822 (88.1%) | 598 [365 - 729] | 5,929 (90.0%) | 598 [365 - 729] | 5,112 (92.8%) | 729 [527 - 729] |
| | 1. South Asian | 1,228 (3.2%) | 594 [426 - 729] | 217 (3.3%) | 594 [426 - 729] | 106 (1.9%) | 729 [426 - 729] |
| | 2. Black | 579 (1.5%) | 668 [426 - 729] | 67 (1.0%) | 668 [426 - 729] | 36 (0.7%) | 729 [426 - 729] |
| | 3. Other/Mixed | 1,238 (3.2%) | 612 [401 - 729] | 195 (3.0%) | 612 [401 - 729] | 151 (2.7%) | 729 [454 - 729] |
| | *Missing* | 1,519 (4.0%) | 556 [365 - 729] | 182 (2.8%) | 556 [365 - 729] | 105 (1.9%) | 729 [588 - 729] |
| **Immunosuppression risk level** | High | 14,856 (38.7%) | 448 [349 - 729] | 2,452 (37.2%) | 448 [349 - 729] | 1,626 (29.5%) | 729 [426 - 729] |
| | Low | 23,530 (61.3%) | 714 [426 - 729] | 4,138 (62.8%) | 714 [426 - 729] | 3,884 (70.5%) | 729 [587 - 729] |
| **Risk category** | 1 (Highest) | 4,360 (11.4%) | 632 [426 - 729] | 449 (6.8%) | 632 [426 - 729] | 950 (17.2%) | 729 [542 - 729] |
| | 2 | 1,046 (2.7%) | 729 [644 - 729] | 132 (2.0%) | 729 [644 - 729] | 223 (4.0%) | 729 [512 - 729] |
| | 3 | 9,450 (24.6%) | 399 [275 - 577] | 1,871 (28.4%) | 399 [275 - 577] | 453 (8.2%) | 520 [408 - 729] |
| | 4 | 12,965 (33.8%) | 494 [182 - 729] | 2,270 (34.4%) | 494 [182 - 729] | 1,793 (32.5%) | 729 [506 - 729] |
| | 5 (Lowest) | 10,565 (27.5%) | 729 [651 - 729] | 1,868 (28.3%) | 729 [651 - 729] | 2,091 (37.9%) | 729 [656 - 729] |
| **Common mental disorders (CMD)** | No | 28,370 (73.9%) | 610 [367 - 729] | 4,798 (72.8%) | 610 [367 - 729] | 4,264 (77.4%) | 729 [535 - 729] |
| | Yes | 10,016 (26.1%) | 567 [365 - 729] | 1,792 (27.2%) | 567 [365 - 729] | 1,246 (22.6%) | 729 [441 - 729] |
| **Severe mental illness (SMI)** | No | 37,980 (98.9%) | 598 [365 - 729] | 6,529 (99.1%) | 598 [365 - 729] | 5,485 (99.5%) | 729 [524 - 729] |
| | Yes | 406 (1.1%) | 546 [414 - 729] | 61 (0.9%) | 546 [414 - 729] | 25 (0.5%) | 729 [426 - 729] |
| **Dementia** | No | 37,734 (98.3%) | 598 [366 - 729] | 6,501 (98.6%) | 598 [366 - 729] | 5,474 (99.3%) | 729 [520 - 729] |
| | Yes | 652 (1.7%) | 583 [365 - 729] | 89 (1.4%) | 583 [365 - 729] | 36 (0.7%) | 690 [464 - 729] |
| **Diabetes Mellitus** | No | 26,392 (68.8%) | 604 [365 - 729] | 4,402 (66.8%) | 604 [365 - 729] | 3,939 (71.5%) | 729 [526 - 729] |
| | Yes | 11,994 (31.2%) | 583 [373 - 729] | 2,188 (33.2%) | 583 [373 - 729] | 1,571 (28.5%) | 729 [510 - 729] |
| **Chronic obstructive pulmonary disease (COPD)** | No | 31,730 (82.7%) | 621 [384 - 729] | 5,384 (81.7%) | 621 [384 - 729] | 4,911 (89.1%) | 729 [524 - 729] |
| | Yes | 6,656 (17.3%) | 530 [322 - 729] | 1,206 (18.3%) | 530 [322 - 729] | 599 (10.9%) | 729 [492 - 729] |
| **Chronic kidney disease (CKD)** | No | 32,153 (83.8%) | 585 [365 - 729] | 5,685 (86.3%) | 585 [365 - 729] | 4,710 (85.5%) | 729 [514 - 729] |
| | Yes | 6,233 (16.2%) | 668 [416 - 729] | 905 (13.7%) | 668 [416 - 729] | 800 (14.5%) | 729 [561 - 729] |
| **Harmful alcohol use** | No | 37,794 (98.5%) | 601 [367 - 729] | 6,492 (98.5%) | 601 [367 - 729] | 5,453 (99.0%) | 729 [523 - 729] |
| | Yes | 592 (1.5%) | 460 [358 - 729] | 98 (1.5%) | 460 [358 - 729] | 57 (1.0%) | 722 [426 - 729] |

| | | | | | | | |
|---|---|---|---|---|---|---|---|
| **Smoking history** | No | 9,964 (26.0%) | 619 [375 - 729] | 1,692 (25.7%) | 619 [375 - 729] | 1,638 (29.7%) | 729 [486 - 729] |
| | Yes | 27,610 (71.9%) | 593 [365 - 729] | 4,800 (72.8%) | 593 [365 - 729] | 3,781 (68.6%) | 729 [528 - 729] |
| | *Missing* | 812 (2.1%) | 569 [368 - 729] | 98 (1.5%) | 569 [368 - 729] | 91 (1.7%) | 729 [586 - 729] |
| **Overweight/obese** | BMI <25 $kg/m^2$ | 11,036 (28.8%) | 620 [396 - 729] | 1,737 (26.4%) | 620 [396 - 729] | 1,669 (30.3%) | 729 [517 - 729] |
| | BMI ≥25 $kg/m^2$ | 22,970 (59.8%) | 589 [365 - 729] | 4,250 (64.5%) | 589 [365 - 729] | 3,403 (61.8%) | 729 [516 - 729] |
| | NA | 4,380 (11.4%) | 588 [365 - 729] | 603 (9.2%) | 588 [365 - 729] | 438 (7.9%) | 729 [532 - 729] |
| **Living alone** | No | 36,616 (95.4%) | 600 [366 - 729] | 6,298 (95.6%) | 600 [366 - 729] | 5,349 (97.1%) | 729 [524 - 729] |
| | Yes | 1,770 (4.6%) | 558 [365 - 729] | 292 (4.4%) | 558 [365 - 729] | 161 (2.9%) | 729 [456 - 729] |
| **Care home residence** | No | 38,038 (99.1%) | 598 [365 - 729] | 6,555 (99.5%) | 598 [365 - 729] | 5,503 (99.9%) | 729 [522 - 729] |
| | Yes | 348 (0.9%) | 554 [415 - 729] | 35 (0.5%) | 554 [415 - 729] | 7 (0.1%) | 426 [426 - 543] |

Supplementary Table S11: Shingrix® vaccine uptake by ethnicity and IMD quintile (n vaccinated/N in ethnicity+IMD quintile subgroup) (% uptake in ethnicity+IMD quintile group).

| | Deprivation (IMD quintile) | | | | |
|---|---|---|---|---|---|
| **Ethnicity** | **1 - Least Deprived** | **2** | **3** | **4** | **5 - Most Deprived** |
| **White** | 3,116/14,146 (22.0%) | 2,737/14,068 (19.5%) | 2,232/12,352 (18.1%) | 1,771/11,208 (15.8%) | 1,368/9,927 (13.8%) |
| **South Asian** | 57/320 (17.8%) | 47/315 (14.9%) | 69/426 (16.2%) | 72/525 (13.7%) | 61/464 (13.1%) |
| **Black** | 12/52 (23.1%) | 5/65 (7.7%) | 26/187 (13.9%) | 34/347 (9.8%) | 28/385 (7.3%) |
| **Other/Mixed** | 51/327 (15.6%) | 58/378 (15.3%) | 69/452 (15.3%) | 84/588 (14.3%) | 62/485 (12.8%) |

IMD, index of multiple deprivation quintiles.

#### Supplementary Figure S1: Entry dates of individuals into the study.

There was a large number of individuals entering at the start of the study, and then subsequent more common entry dates when people become eligible on their birthday (set as 1st July for all participants due to data only including year of birth).

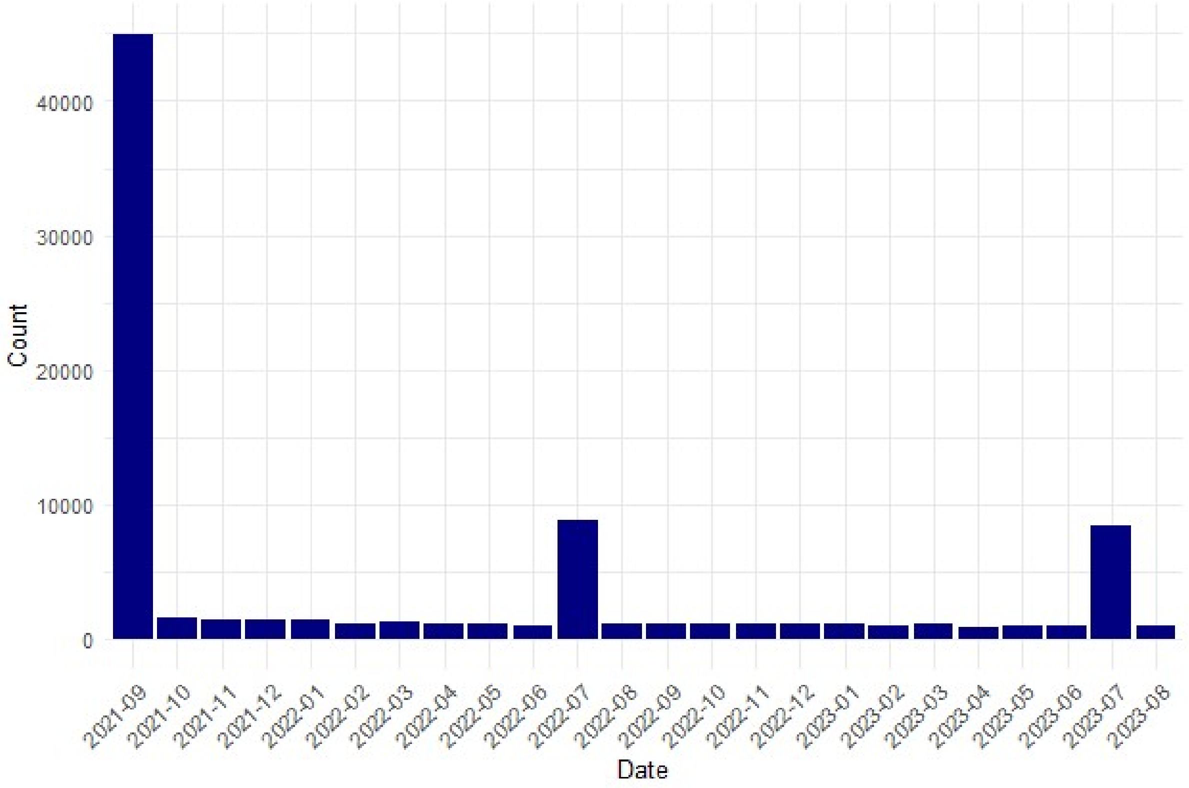

### Separate file: STROBE RECORD checklist

| | Ite m No. | STROBE items | Location in manuscript where items are reported | RECORD items | Location in manuscript where items are reported |
|---|---|---|---|---|---|
| **Title and abstract** | | | | | |
| | 1 | (a) Indicate the study's design with a commonly used term in the title or the abstract (b) Provide in the abstract an informative and balanced summary of what was done and what was found | Title and abstract | RECORD 1.1: The type of data used should be specified in the title or abstract. When possible, the name of the databases used should be included.<br><br>RECORD 1.2: If applicable, the geographic region and timeframe within which the study took place should be reported in the title or abstract.<br><br>RECORD 1.3: If linkage between databases was conducted for the study, this should be clearly stated in the title or abstract. | Title and abstract<br><br>Abstract<br><br>Not applicable |
| **Introduction** | | | | | |
| Background rationale | 2 | Explain the scientific background and rationale for the investigation being reported | Abstract and introduction | | |
| Objectives | 3 | State specific objectives, including any prespecified hypotheses | Introduction | | |
| **Methods** | | | | | |
| Study Design | 4 | Present key elements of study design early in the paper | Methods | | |
| Setting | 5 | Describe the setting, locations, and relevant dates, including periods of recruitment, exposure, follow-up, and data collection | Methods (Data sources, Study population) | | |
| Participants | 6 | *(a) Cohort study* - Give the eligibility criteria, and the sources and methods of selection of participants. Describe methods of follow-up *Case-control study* - Give the eligibility criteria, and the sources and methods of case ascertainment and control selection. Give the rationale for the choice of cases and controls | Methods (Study population, Outcomes), Supplementary Tables S2, S3, S4. | RECORD 6.1: The methods of study population selection (such as codes or algorithms used to identify subjects) should be listed in detail. If this is not possible, an explanation should be provided.<br><br>RECORD 6.2: Any validation studies of the codes or algorithms used to select the population should be referenced. If validation was conducted for this study and not published elsewhere, detailed methods and results should be provided. | Methods (Study population, Outcomes). Supplementary text S1, Supplementary Table S1, S2, S3.<br><br>For the operationalisation of the immunosuppression definitions, we relied on the code lists and algorithms developed by the University of Nottingham Primary Care Information Services (PRIMIS) in collaboration with the UK Health Security Agency |

| | | | | | |
|---|---|---|---|---|---|
| | | *Cross-sectional study* - Give the eligibility criteria, and the sources and methods of selection of participants<br><br>*(b) Cohort study* - For matched studies, give matching criteria and number of exposed and unexposed<br>*Case-control study* - For matched studies, give matching criteria and the number of controls per case | | RECORD 6.3: If the study involved linkage of databases, consider use of a flow diagram or other graphical display to demonstrate the data linkage process, including the number of individuals with linked data at each stage. | (UKHSA). PRIMIS have developed comprehensive and reliable code lists, which were specifically designed to identify individuals eligible for vaccine programmes, including those defined as immunosuppressed in the Green Book criteria. All definitions were reviewed by people with clinical experience, and our code list was formulated to match the guidelines. The code list can be found as specified in GitHub repository doi:10.5281/zenodo.15351553<br><br>No linkage was used to justify participation. |
| Variables | 7 | Clearly define all outcomes, exposures, predictors, potential confounders, and effect modifiers. Give diagnostic criteria, if applicable. | Methods (Outcomes, Exposures and other variables) | RECORD 7.1: A complete list of codes and algorithms used to classify exposures, outcomes, confounders, and effect modifiers should be provided. If these cannot be reported, an explanation should be provided. | Supplementary Materials and table of variables can be found in GitHub repository doi:10.5281/zenodo.15351553. |
| Data sources/ measurement | 8 | For each variable of interest, give sources of data and details of methods of assessment (measurement). Describe comparability of assessment methods if there is more than one group | Methods (Data sources, Exposures and other variables). | | |
| Bias | 9 | Describe any efforts to address potential sources of bias | Methods, Results, Supplementary Table S4 and S5, | | |
| Study size | 10 | Explain how the study size was arrived at | All individuals meeting the eligibility criteria were included in this cohort study (Study flowchart: Figure 1) | | |
| Quantitative variables | 11 | Explain how quantitative variables were handled in the analyses. If applicable, describe which groupings were chosen, and why | Methods (Exposures and other variables, Statistical analysis), Supplementary Table S4 and S5) | | |

| | | | | | |
|---|---|---|---|---|---|
| Statistical methods | 12 | (a) Describe all statistical methods, including those used to control for confounding<br>(b) Describe any methods used to examine subgroups and interactions<br>(c) Explain how missing data were addressed<br>(d) *Cohort study* - If applicable, explain how loss to follow-up was addressed<br>*Case-control study* - If applicable, explain how matching of cases and controls was addressed<br>*Cross-sectional study* - If applicable, describe analytical methods taking account of sampling strategy<br>(e) Describe any sensitivity analyses | Statistical analyses and Supplementary Tables S4 and S5.<br><br>Supplementary Tables S7, S9, S10, S11<br><br>Statistical analysis<br><br>Not applicable<br><br>Supplementary Tables S5. | | |
| Data access and cleaning methods | | .. | | RECORD 12.1: Authors should describe the extent to which the investigators had access to the database population used to create the study population.<br><br>RECORD 12.2: Authors should provide information on the data cleaning methods used in the study. | Study population. Deidentified patients were identified as being in the study period and having immunosuppression codes. Their medical records were then extracted.<br><br>R codes can be found in GitHub repository doi:10.5281/zenodo.15351553. |
| Linkage | | .. | | RECORD 12.3: State whether the study included person-level, institutional-level, or other data linkage across two or more databases. The methods of linkage and methods of linkage quality evaluation should be provided. | Methods: Data source |
| **Results** | | | | | |
| Participants | 13 | (a) Report the numbers of individuals at each stage of the study (*e.g.*, numbers potentially eligible, examined for eligibility, confirmed eligible, included in the | Reported in results and Figure 1 presents the study flow chart. | RECORD 13.1: Describe in detail the selection of the persons included in the study (*i.e.,* study population selection) including filtering based on data quality, data availability and linkage. The selection of included persons can be described in the text and/or by means of the study flow diagram. | Study population, Figure 1, Supplementary Materials, Supplementary Tables S1, S2, S3. |

| | | | |
|---|---|---|---|
| | | study, completing follow-up, and analysed)<br>(b) Give reasons for non-participation at each stage.<br>(c) Consider use of a flow diagram | |
| Descriptive data | 14 | (a) Give characteristics of study participants (*e.g.*, demographic, clinical, social) and information on exposures and potential confounders<br>(b) Indicate the number of participants with missing data for each variable of interest<br>(c) *Cohort study* - summarise follow-up time (*e.g.*, average and total amount) | Table 2 provides participants' characteristics and information on missing data, as well as information on follow-up time. |
| Outcome data | 15 | *Cohort study* - Report numbers of outcome events or summary measures over time<br>*Case-control study* - Report numbers in each exposure category, or summary measures of exposure<br>*Cross-sectional study* - Report numbers of outcome events or summary measures | Results and Table 2 provide outcome data and counts. |
| Main results | 16 | (a) Give unadjusted estimates and, if applicable, confounder-adjusted estimates and their precision (e.g., 95% confidence interval). Make clear which confounders were adjusted for and why they were included<br>(b) Report category boundaries when continuous variables were categorized | Supplementary Table S6 provides unadjusted odds ratio for uptake (as described in Supplementary Table S4). Figure 2 provides adjusted odds ratios with 95% confidence intervals and provides information for the variables that were adjusted for. |

| | | | | | |
|---|---|---|---|---|---|
| | | (c) If relevant, consider translating estimates of relative risk into absolute risk for a meaningful time period | | | |
| Other analyses | 17 | Report other analyses done—e.g., analyses of subgroups and interactions, and sensitivity analyses | All sensitivity analyses are summarised in Supplementary Table S5 with results shown in Supplementary Table S7. Further granularity for risk level and ethnicity are shown in Supplementary Table S7 and S9, respectively. Hierarchical modelling is shown in Supplementary Table S4. Interaction of ethnicity and IMD are shown in Supplementary Table S11. | | |
| **Discussion** | | | | | |
| Key results | 18 | Summarise key results with reference to study objectives | Discussion (Summary) | | |
| Limitations | 19 | Discuss limitations of the study, taking into account sources of potential bias or imprecision. Discuss both direction and magnitude of any potential bias | Discussion (Strengths and Limitations) | RECORD 19.1: Discuss the implications of using data that were not created or collected to answer the specific research question(s). Include discussion of misclassification bias, unmeasured confounding, missing data, and changing eligibility over time, as they pertain to the study being reported. | Discussed (Strengths and limitations). |
| Interpretation | 20 | Give a cautious overall interpretation of results considering objectives, limitations, multiplicity of analyses, results from similar studies, and other relevant evidence | Discussion | | |
| Generalisability | 21 | Discuss the generalisability (external validity) of the study results | Discussion (Possible explanations and implications for policy and practice) | | |
| **Other Information** | | | | | |
| Funding | 22 | Give the source of funding and the role of the funders for the present study and, if applicable, for the original study on which the present article is based | Competing interest statement; and Funding and statement of independence of researchers from funders | | |

| | | | | | |
|---|---|---|---|---|---|
| Accessibility of protocol, raw data, and programming code | | | | RECORD 22.1: Authors should provide information on how to access any supplemental information such as the study protocol, raw data, or programming code. | Online submission (research protocol) & Github repository for code and codelists (doi:10.5281/zenodo.15351553). Raw data will not be made available due to data protection. |
